# Description of Patient Characteristics and Clinical and Economic Outcomes of Orthopedic Sutures Used for Rotator Cuff Repair

**DOI:** 10.1101/2024.11.21.24317728

**Authors:** Katherine Corso, Caroline Smith, Biju Varughese, James Wood, Jill Ruppenkamp, Matthew Putnam

## Abstract

**Background:** A better understanding of real-world clinical and economic outcomes associated with orthopedic sutures used for rotator cuff repair is needed.

**Research Design and Methods:** A retrospective, descriptive cohort study using Premier Healthcare Database hospital-based data evaluated patients treated with Dynacord™ or FiberWire^®^ for rotator cuff repair between January 2017-February 2022. Baseline patient characteristics and twelve-month outcomes were assessed. Each suture cohort was analyzed separately. No comparative analyses were performed in this study.

**Results:** Baseline demographics of 1 074 patients treated with Dynacord™ and 19 899 treated with FiberWire^®^, respectively, were mean age 59.4, 59.9 years; 59.4% and 58.5% male. All-cause twelve-month hospital revisits were observed in 31.4% and 42.4%, and the incidence proportion with shoulder-related hospital revisits was 12.6% and 14.0%, respectively. The incidence of twelve-month rotator cuff re-repair was 1.2% for Dynacord™ and 1.8% for FiberWire^®^. Dynacord™ twelve-month all-cause revisit costs (standard deviation [SD]) were $1 649 ($5 870) and $2 230 ($7 204) for FiberWire^®^. Twelve-month complications, device removals, and manipulations were ≤1% for both cohorts.

**Conclusions:** This study contributes clinical and economic real-world evidence on two types of orthopedic sutures used for rotator cuff repair.

## Introduction

Shoulder pain is highly prevalent in older populations and may accelerate functional decline.^1^ Rotator cuff tears are the most common musculoskeletal shoulder derangement, affecting at least 10% of people over the age of 60 in the United States (US).^2^ Operative intervention for rotator cuff repair is intended to reduce pain and improve strength and range of motion.^1^ It is estimated that 570 000 rotator cuff repairs were performed in the US in 2023.^3^ Over time, the comorbidity profile of patients undergoing rotator cuff repair is becoming more complex with greater prevalence of numerous comorbidities, including hypertension, peripheral vascular disease, and chronic pulmonary disease.^4^

Rotator cuff repairs continue to fail at an alarming rate, with re-tear rates for large rotator cuff repairs ranging from 20-40% and re-tear rates for massive and significantly retracted rotator cuff tears up to 94%.^5^ Repair of a recurrent rotator cuff tear is more difficult due to shorter tendons and excess implant at the footprint.^6^ A solution that maintains continuous compression throughout the healing period, provides a more stable footprint, and reduces the risk of suture pull-through is needed.^7-14^ This may help reduce the risk of rotator cuff repair failure, and in turn reduce the economic burden of rotator cuff repair failure to the hospital and healthcare system.

Dynacord™ (DePuy Synthes, Raynham, MA, USA) is a medical suture for shoulder rotator cuff repair that consists of three layers: 1) an outer braided sheath made of ultra-high molecular weight polyethylene fibers (UHMWPE) and an optional polyester tracer; 2) an inner sheath made of braided polyester fibers; and 3) a clear silicone layer co-extruded over a salt-infused silicone core.^15^ FiberWire^®^ (Arthrex, Inc., Naples, FL, USA) is a multi-strand, long chain UHMWPE core with a braided jacket of polyester and UHMWPE.^16^

There is a lack of published real-world evidence on the performance and safety of Dynacord™ and FiberWire^®^ in patients with rotator cuff repair. A better understanding of the real-world outcomes and healthcare costs associated with rotator cuff repair with Dynacord™ and FiberWire^®^ procedures may assist healthcare providers and payers in identifying opportunities for improving rotator cuff patient quality of care while simultaneously reducing overall healthcare costs. The objectives of this study were to describe only and not compare the clinical and economic burden of two types of orthopedic sutures, Dynacord™ and FiberWire^®^, in the year following rotator cuff repair.

## Patients and Methods

### Data Source

This retrospective, descriptive, observational study using real-world data identified patients undergoing rotator cuff repair in the *Premier Healthcare Database* (PHD) between January 2017 and February 2022. The PHD contains complete standardized clinical coding, including diagnosis, procedure, and hospital-prescribed medications from more than 25% of all hospital admissions throughout the US (>1 164 hospitals and hospital systems). Premier, Inc. collects data from participating hospitals in its health care alliance. Although the database excludes federally funded hospitals (e.g., Veterans Affairs), the hospitals included are nationally representative based on bed size, geographic region, location (urban/rural) and teaching hospital status. The database contains a date-stamped log of all billed items by the cost-accounting department which includes medications; laboratory, diagnostic, and therapeutic services; and primary and secondary diagnoses and procedures for each patient’s hospitalization. Identifier-linked enrollment files provide demographic and payer information. Detailed service level information for each hospital day is recorded; this includes details on medication and devices received and associated hospital charges and costs of each item.

The use of the PHD was reviewed by the New England Institutional Review Board (IRB) and was determined to be exempt from broad IRB approval, as this research project did not involve human subject research and used data from an anonymous, de-identified, administrative claims database compliant with the Health Insurance Portability and Accountability Act of 1996.

### Patient Population

Patients were considered for inclusion in the study if they met all of the following inclusion criteria: a billing charge description for Dynacord™ or FiberWire^®^ from January 2017 and February 2022; a Current Procedural Terminology (CPT) code or International Classification of Diseases, Tenth Revision (ICD-10) procedure code indicative of initial rotator cuff repair; a diagnosis for rotator cuff degenerative or traumatic condition; an elective procedure; age ≥18 years old; and continuous enrollment 3 months prior to and 12 months after the index date. The index date was the admission date for the rotator cuff repair surgery performed in the inpatient setting, or the service day for same day/outpatient procedures. Patients meeting any of the following criteria were excluded from the study: evidence of revision rotator cuff procedure at index; concurrent diagnosis of infection of the shoulder/upper arm or shoulder fracture; fracture of any anatomy in the 3 months prior to index; two or more suture types; unknown patient age or sex; death during the index admission; or bilateral rotator cuff repairs at index or in follow-up. CPT and ICD-10 codes used for patient identification are located in the **Supplemental File**. Dynacord™ was launched in the marketplace in the year 2018 and FiberWire^®^ was launched in year 2001; thus the study years (2017 to 2022) were chosen to have a study start date in the year prior to adoption of Dynacord™.

## Study Measures

### Baseline Characteristics

Patient demographic and clinical characteristics that were evaluated included age, gender, race, marital status, payer category (Medicare, Medicaid, or commercial health insurance), and patient comorbidities. Baseline comorbidity was assessed using the Elixhauser Comorbidity Score. The Elixhauser Comorbidity score is an aggregate measure of comorbidity created by using 31 dimensions associated with chronic disease (e.g., heart disease, cancer) and overall health conditions. Prior research has shown that increasing Elixhauser Comorbidity Scores are associated with increased healthcare utilization and greater risk of mortality.^17, 18^ Information regarding tobacco use at time of rotator cuff repair, the setting of care (inpatient, outpatient, or other), the year of surgery (2017-2022), clinical status (trauma, degenerative, or both), approach (arthroscopic, open, or both), and laterality (left, right, or unknown) was also collected. Provider characteristics that were evaluated included whether the procedure was performed in an urban or rural setting, whether the hospital was a teaching hospital, the hospital Census region, hospital bed size, and annual procedure volume.

### Hospital Clinical Outcomes, Resource Utilization, and Costs

Study outcomes were analyzed for 12 months following index rotator cuff repair. The incidence proportions and hospital costs of all-cause and shoulder-related hospital revisits, re-repair, and complications in the year following rotator cuff repair were analyzed separately for Dynacord™ and FiberWire^®^. For each cohort in this study, the incidence proportions were calculated by dividing the number of patients at the start of the study by the number of events that occurred, and were expressed as a percent per each outcome. Shoulder-related hospital revisits were defined as a hospital revisit for a shoulder diagnosis or procedure. Re-repairs were defined as return for a second rotator cuff repair. Complication events collected were tissue cuts, mechanical complications, foreign body reaction, and infections. Shoulder-related hospital revisits, re-repairs, and complications were collected using CPT or ICD codes which can be found in the **Supplemental File**.

### Statistical Analyses

Descriptive statistics were performed to describe all variables in the dataset for patients who received each of the two devices separately. No statistical comparisons were performed in this study because the intent of this study was not to compare the devices, but rather to describe their burden. Continuous variables were presented using mean and standard deviations while categorical or binary variables were presented as frequencies and percentages. Clinical outcomes, healthcare resource utilization outcomes, and costs were presented as percentages with 95% confidence intervals (CIs). All costs were inflated to 2022 US dollars using the Bureau of Labor Statistics (BLS) consumer price-index.^19^

## Results

### Patient Selection

A total of 1 074 patients from the PHD received Dynacord™ and 19 899 patients received FiberWire^®^ between January 2017 and February 2022.

### Patient Baseline Demographic and Clinical Characteristics

Baseline demographic and clinical characteristics for patients with Dynacord™ and FiberWire^®^ rotator cuff repair are presented in **Table 1**. Mean (SD) age was 59.4 (9.3) years old for Dynacord™ and 59.9 (10.0) years old for FiberWire^®^, 59.4% of Dynacord™ and 58.5% of FiberWire^®^ patients were male, the majority of patients were White (Dynacord™ 88.5% and FiberWire^®^ 81.3%), and most patients were married (Dynacord™ 70.2% and FiberWire^®^ 65.2%). Most patients had private commercial health insurance (Dynacord™ 45.2% and FiberWire^®^ 44.0%) or Medicare (Dynacord™ 29.7% and FiberWire^®^ 33.9%).

**Table 1.** Baseline patient demographic characteristics by suture (<u>no statistical adjustment</u> <u>for differences in cohorts conducted</u>)

| Patient Characteristic | Dynacord™ | FiberWire® |
| --- | --- | --- |
| **N** | 1 074 | 19 899 |
| **Age in years, mean (SD)** |  |  |
| Mean (SD) | 59.4 (9.3) | 59.9 (10.0) |
| **Age in years, n (%)** |  |  |
| 18-34 | 5 (0.5) | 216 (1.1) |
| 35-44 | 57 (5.3) | 1 120 (5.6) |
| 45-54 | 248 (23.1) | 4 296 (21.6) |
| 55-64 | 443 (41.2) | 7 539 (37.9) |
| 65-74 | 272 (25.3) | 5 497 (27.6) |
| ≥75 | 49 (4.6) | 1 231 (6.2) |
| **Gender, n (%)** |  |  |
| Males | 638 (59.4) | 11 646 (58.5) |
| **Race, n (%)** |  |  |
| Asian | 5 (0.5) | 607 (3.1) |
| Black | 51 (4.7) | 1 786 (9.0) |
| Other/Unknown | 68 (6.3) | 1 334 (6.7) |
| White | 950 (88.5) | 16 172 (81.3) |
| **Marital Status, n (%)** |  |  |
| Married | 754 (70.2) | 12 983 (65.2) |
| Other | 7 (0.7) | 723 (3.6) |
| Single | 309 (28.8) | 6 124 (30.8) |
| Unknown | 4 (0.4) | 69 (0.3) |
| **Payer category, n (%)** |  |  |
| Commercial | 485 (45.2) | 8 748 (44.0) |
| Medicaid | 73 (6.8) | 1 307 (6.6) |
| Medicare | 319 (29.7) | 6 736 (33.9) |
| Other | 197 (18.3) | 3 108 (15.6) |

SD, standard deviation.

Few patients were admitted as hospital inpatients (Dynacord™ 0.5% and FiberWire^®^ 1.9%) (**Table 2**). Most patients who were treated with Dynacord™ had their rotator cuff repair in 2021 (∼40%), and for FiberWire^®^ most had their repair in the earlier years of the study, 2017 to 2019 (∼22% per year). Most patients had degenerative disease (Dynacord™ 88.1% and FiberWire^®^ 82.2%), arthroscopic approach (Dynacord™ 98.2% and FiberWire^®^ 83.8%), and right laterality (Dynacord™ 61.7% and FiberWire^®^ 62.0%). Mean (SD) Elixhauser Comorbidity Index scores were 0.94 (1.24) for patients who were repaired with Dynacord™ and 1.28 (1.32) for patients who were repaired with FiberWire^®^. Approximately one-quarter of patients were smokers (Dynacord™ 24.8% and FiberWire^®^ 28.9%).

**Table 2.**
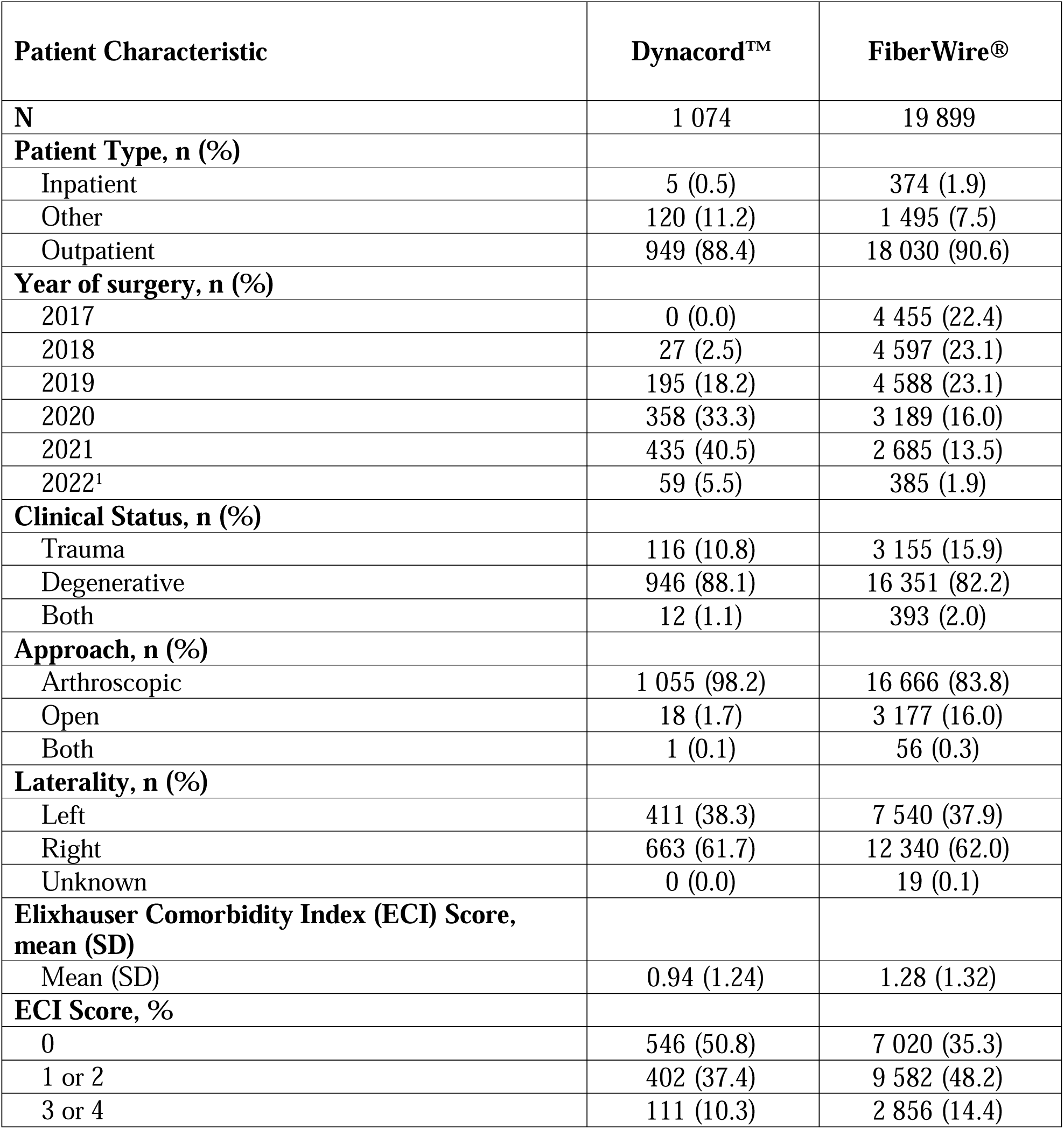
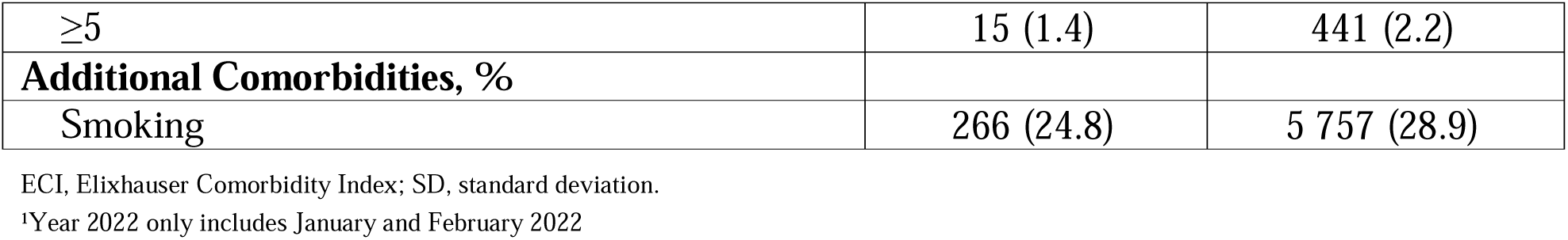
Baseline patient clinical characteristics by suture (<u>no statistical adjustment for</u> <u>differences in cohorts conducted</u>)

| Patient Characteristic | Dynacord™ | FiberWire® |
| --- | --- | --- |
| **N** | 1 074 | 19 899 |
| **Patient Type, n (%)** |  |  |
| Inpatient | 5 (0.5) | 374 (1.9) |
| Other | 120 (11.2) | 1 495 (7.5) |
| Outpatient | 949 (88.4) | 18 030 (90.6) |
| **Year of surgery, n (%)** |  |  |
| 2017 | 0 (0.0) | 4 455 (22.4) |
| 2018 | 27 (2.5) | 4 597 (23.1) |
| 2019 | 195 (18.2) | 4 588 (23.1) |
| 2020 | 358 (33.3) | 3 189 (16.0) |
| 2021 | 435 (40.5) | 2 685 (13.5) |
| 2022<sup>1</sup> | 59 (5.5) | 385 (1.9) |
| **Clinical Status, n (%)** |  |  |
| Trauma | 116 (10.8) | 3 155 (15.9) |
| Degenerative | 946 (88.1) | 16 351 (82.2) |
| Both | 12 (1.1) | 393 (2.0) |
| **Approach, n (%)** |  |  |
| Arthroscopic | 1 055 (98.2) | 16 666 (83.8) |
| Open | 18 (1.7) | 3 177 (16.0) |
| Both | 1 (0.1) | 56 (0.3) |
| **Laterality, n (%)** |  |  |
| Left | 411 (38.3) | 7 540 (37.9) |
| Right | 663 (61.7) | 12 340 (62.0) |
| Unknown | 0 (0.0) | 19 (0.1) |
| **Elixhauser Comorbidity Index (ECI) Score, mean (SD)** |  |  |
| Mean (SD) | 0.94 (1.24) | 1.28 (1.32) |
| **ECI Score, %** |  |  |
| 0 | 546 (50.8) | 7 020 (35.3) |
| 1 or 2 | 402 (37.4) | 9 582 (48.2) |
| 3 or 4 | 111 (10.3) | 2 856 (14.4) |
| ≥5 | 15 (1.4) | 441 (2.2) |
| **Additional Comorbidities, %** |  |  |
| Smoking | 266 (24.8) | 5 757 (28.9) |

ECI, Elixhauser Comorbidity Index; SD, standard deviation.
<sup>1</sup>Year 2022 only includes January and February 2022

### Provider Baseline Characteristics

Baseline provider characteristics are presented in **Table 3**. Most patients in both cohorts were admitted to urban hospitals (Dynacord™ 96.6% and FiberWire^®^ 87.4%). Most patients who received Dynacord™ had their repair in a teaching hospital and patients receiving FiberWire^®^ had their repair in a nonteaching hospital (Dynacord™, 58.8% and FiberWire^®^, 65.6%). Dynacord™ cases were predominantly conducted in Northeast (49.8%) and South (26.4%), and majority of FiberWire^®^ cases were in the South (38.1%) and Midwest (31.1%). Patients with Dynacord™ came from hospitals with a mean annual procedure volume of 176.0 [SD 106.2] rotator cuff cases and for patients with FiberWire^®^ mean annual procedure volume was 160.0 [SD 131.6] cases.

**Table 3.** Baseline provider characteristics by suture (no statistical adjustment for differences in cohorts conducted)

| **Provider Characteristic** | **Dynacord™** | **FiberWire®** |
| --- | --- | --- |
| **N** | 1 074 | 19 899 |
| **Urban or Rural, n (%)** |  |  |
| Urban | 1 038 (96.6) | 17 396 (87.4) |
| **Teaching, n (%)** |  |  |
| Yes | 632 (58.8) | 6 854 (34.4) |
| **Division, n (%)** |  |  |
| Midwest | 239 (22.3) | 6 182 (31.1) |
| Northeast | 535 (49.8) | 2 614 (13.1) |
| South | 283 (26.4) | 7 577 (38.1) |
| West | 17 (1.6) | 3 526 (17.7) |
| **Annual Procedure Volume, mean (SD)** |  |  |
| Mean (SD) | 176.0 (106.2) | 160.4 (131.6) |
| **Hospital Bed Size, n (%)** |  |  |
| 0-99 | 112 (10.4) | 3 527 (17.7) |
| 100-199 | 147 (13.7) | 3 756 (18.9) |
| 200-299 | 580 (54.0) | 4 354 (21.9) |
| 300-399 | 60 (5.6) | 4 174 (21.0) |
| 400-499 | 114 (10.6) | 931 (4.7) |
| ≥500 | 61 (5.7) | 3 157 (15.9) |

SD, standard deviation.

### Hospital-Based Index Healthcare Utilization

The index healthcare utilization for procedures performed with each suture is summarized in **Table 4**. The index hospital mean (SD) cost of surgery with each suture was $10 357 ($5 462) for Dynacord™ and $9 166 ($5 356) for FiberWire^®^. Mean (SD) surgery times were 140.0 (37.6) minutes for Dynacord™ and 144.0 (84.7) minutes for FiberWire^®^. Most patients were discharged home (Dynacord™ 99.4% and FiberWire^®^ 98.7%).

**Table 4.** Index healthcare utilization by suture (no statistical adjustment for differences in cohorts conducted)

| Index measure | Dynacord™ | FiberWire® |
| --- | --- | --- |
| **N** | 1 074 | 19 899 |
| **Index Cost, Adjusted, mean (SD)** | \$10 357 (\$5 462) | \$9 166 (\$5 356) |
| **Surgery Time in minutes, mean (SD)\*** | 140.0 (37.6) | 144.0 (84.7) |
| **Discharge Status, n (%)** |  |  |
| Discharged to HHO | 3 (0.3) | 109 (0.5) |
| Home | 1 068 (99.4) | 19 643 (98.7) |
| SNF & Other | 3 (0.3) | 147 (0.7) |

HHO, home health organization; SD, standard deviation; SNF, skilled nursing facility.
\*Because of missing surgery time in the database, estimates are for 1,066 Dynacord™ patients and 18,444 FiberWire® patients.

### Twelve-Month Outcomes and Associated Costs

Twelve-month all-cause hospital revisits were 31.4% for Dynacord™ patients and 42.4% for FiberWire^®^ (**Figure 1**). The incidence of patients with twelve-month shoulder-related hospital revisits was 12.6% for Dynacord™ and 14.0% for FiberWire^®^ (**Figure 2**). The incidences of twelve-month rotator cuff re-repair were 1.2% and 1.8% for repairs performed with Dynacord™ and FiberWire^®^, respectively (**Figure 3**). Incidences of twelve-month complications, device removals, and manipulations were ≤1% for both cohorts (**Table 5**).

**Figure 1.**
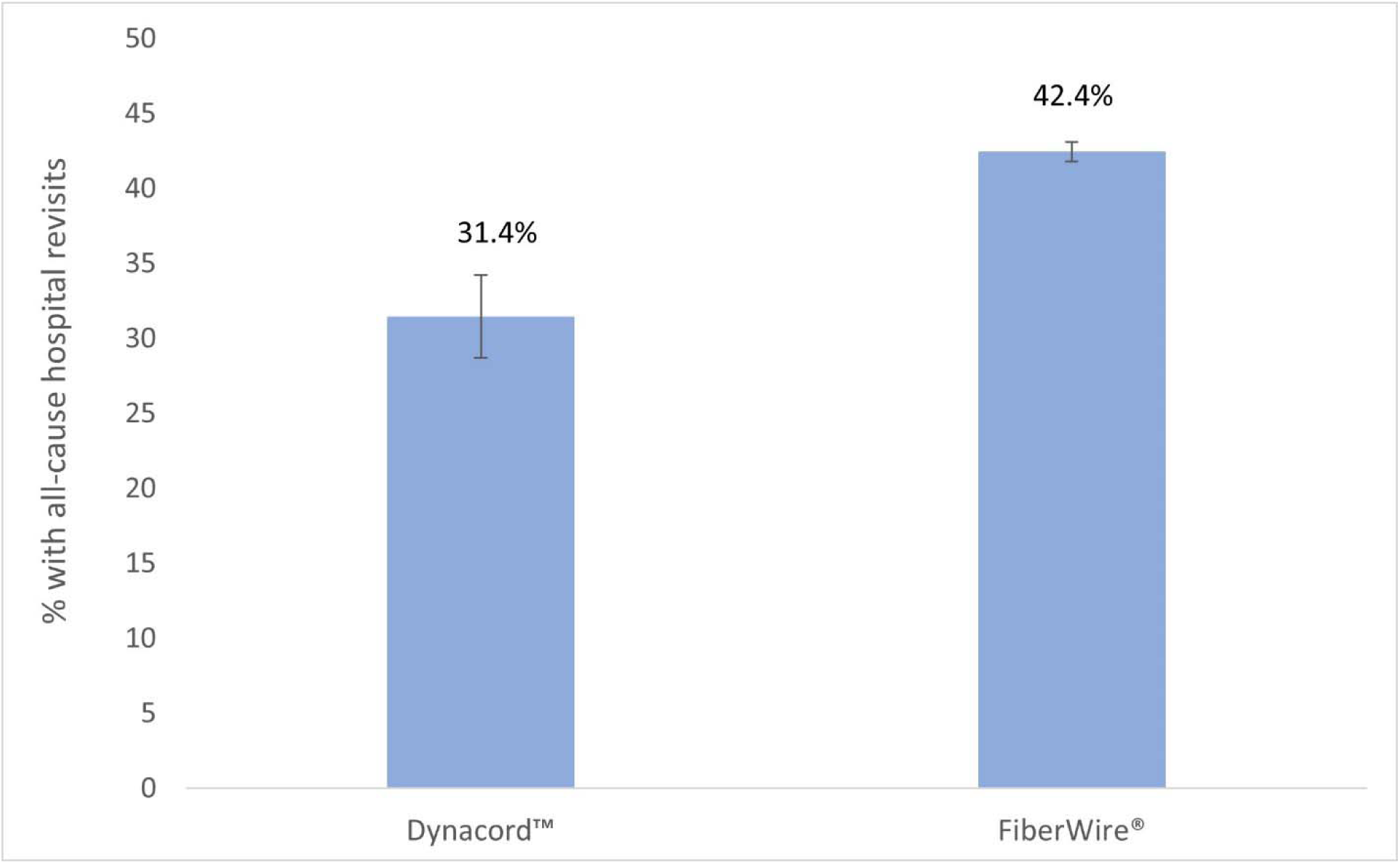
Incidence (% and 95% CI) of 12-month all-cause hospital revisits by suture (<u>no</u> <u>statistical adjustment for differences in cohorts conducted</u>)

**Figure 2.**
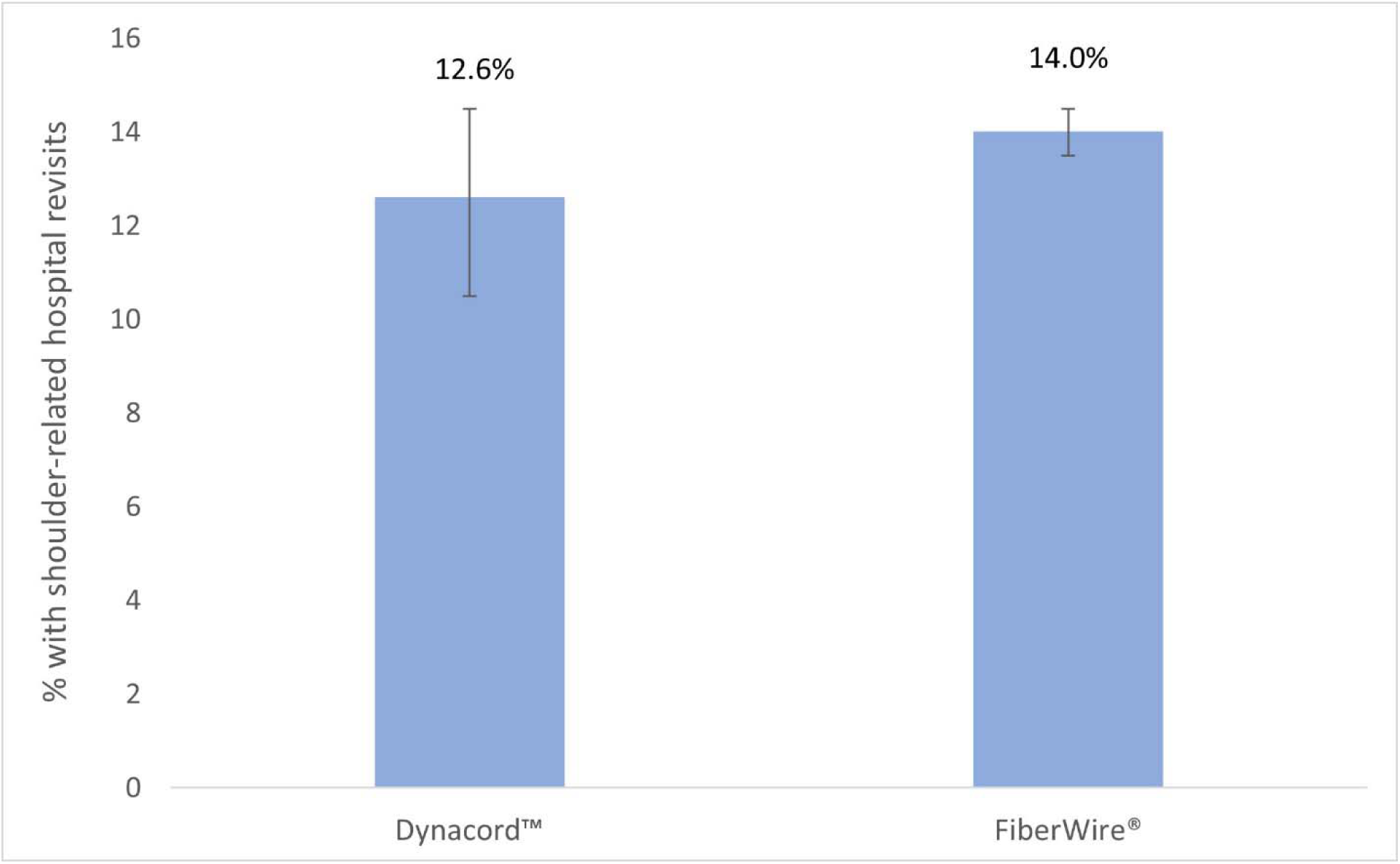
Incidence (% and 95% CI) of 12-month shoulder-related hospital revisits by suture (<u>no statistical adjustment for differences in cohorts conducted</u>)

**Figure 3.**
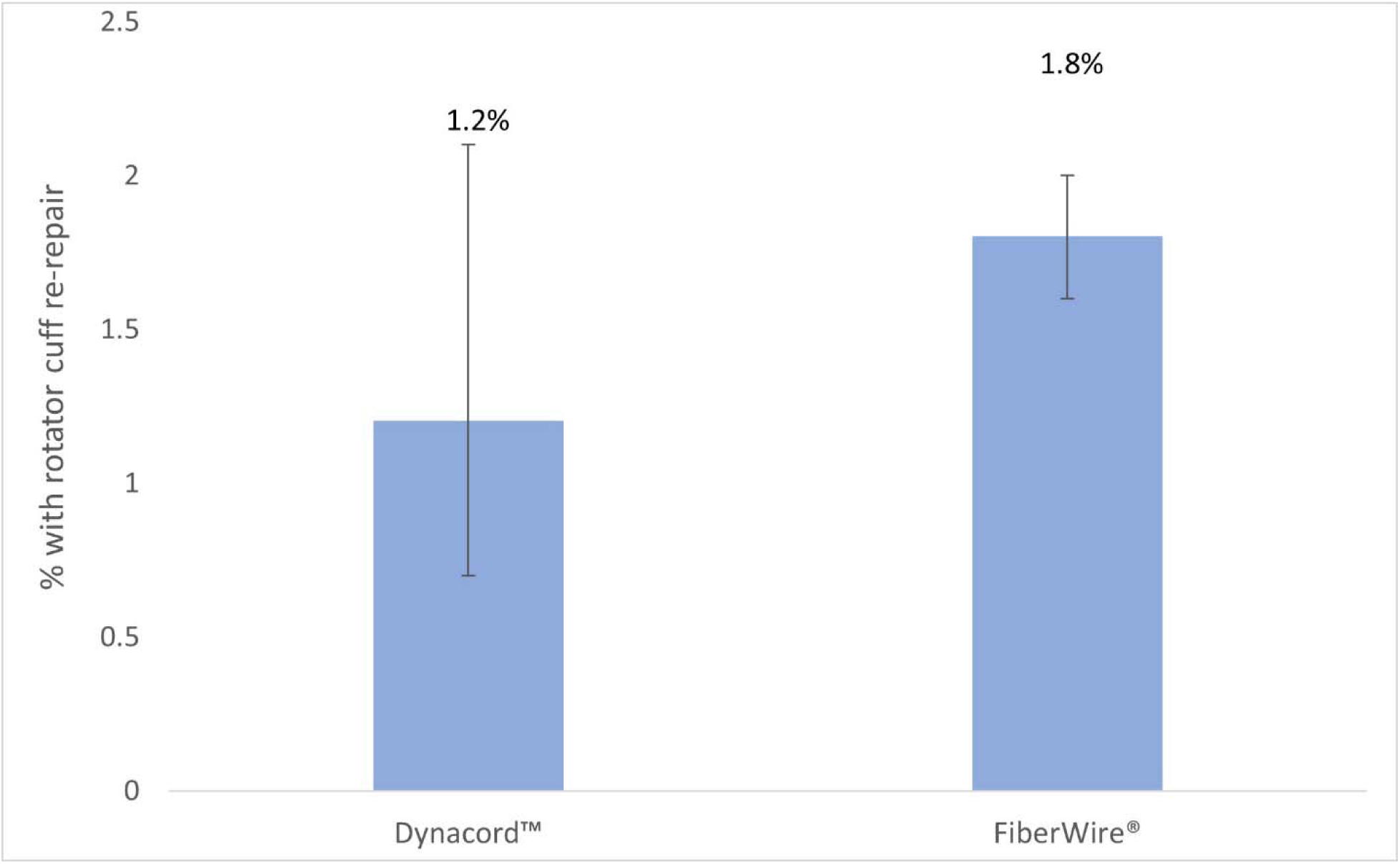
Incidence (% and 95% CI) of 12-month rotator cuff re-repair by suture (<u>no</u> <u>statistical adjustment for differences in cohorts conducted</u>)

**Table 5.**
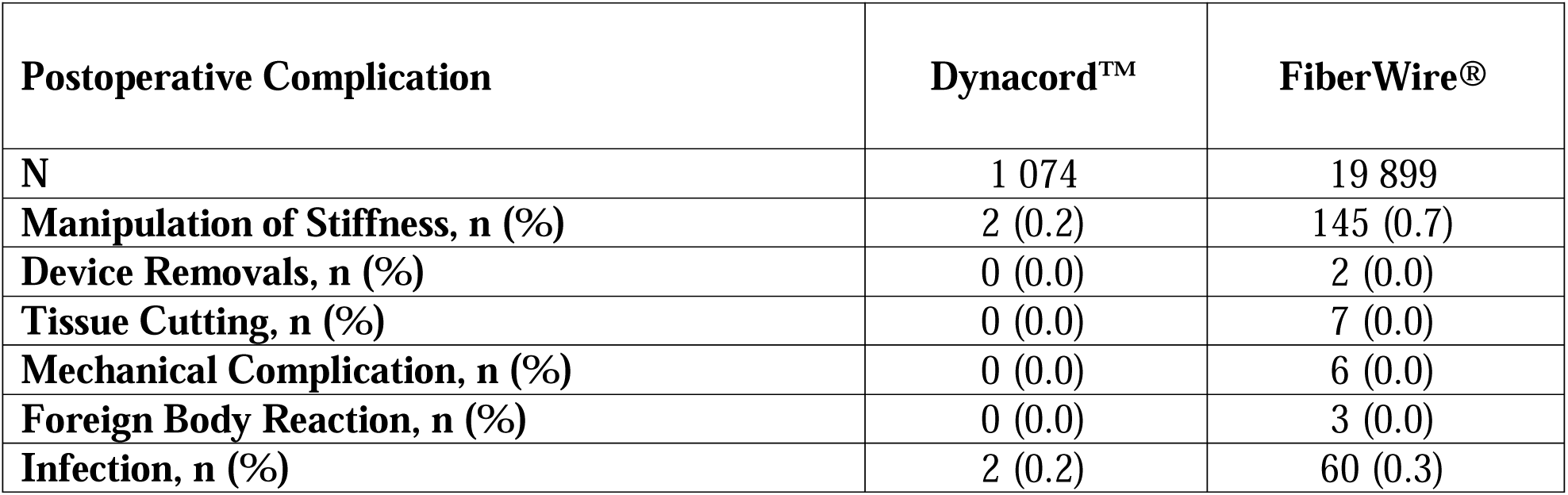
Twelve-month postoperative complications (no statistical adjustment for differences in cohorts conducted)

| **Postoperative Complication** | **Dynacord™** | **FiberWire®** |
| --- | --- | --- |
| **N** | 1 074 | 19 899 |
| **Manipulation of Stiffness, n (%)** | 2 (0.2) | 145 (0.7) |
| **Device Removals, n (%)** | 0 (0.0) | 2 (0.0) |
| **Tissue Cutting, n (%)** | 0 (0.0) | 7 (0.0) |
| **Mechanical Complication, n (%)** | 0 (0.0) | 6 (0.0) |
| **Foreign Body Reaction, n (%)** | 0 (0.0) | 3 (0.0) |
| **Infection, n (%)** | 2 (0.2) | 60 (0.3) |

Patients with Dynacord™ sutures had mean (SD) twelve-month all-cause revisit costs of $1 649 ($5 870) and FiberWire^®^ mean (SD) revisit costs were $2 230 ($7 204) (**Figure 4**). The mean (SD) total twelve-month healthcare costs (index costs + twelve-month revisit costs) was $12 006 [$8 570] for Dynacord™ and $11 395 [$9 089] for FiberWire^®^.

**Figure 4.**
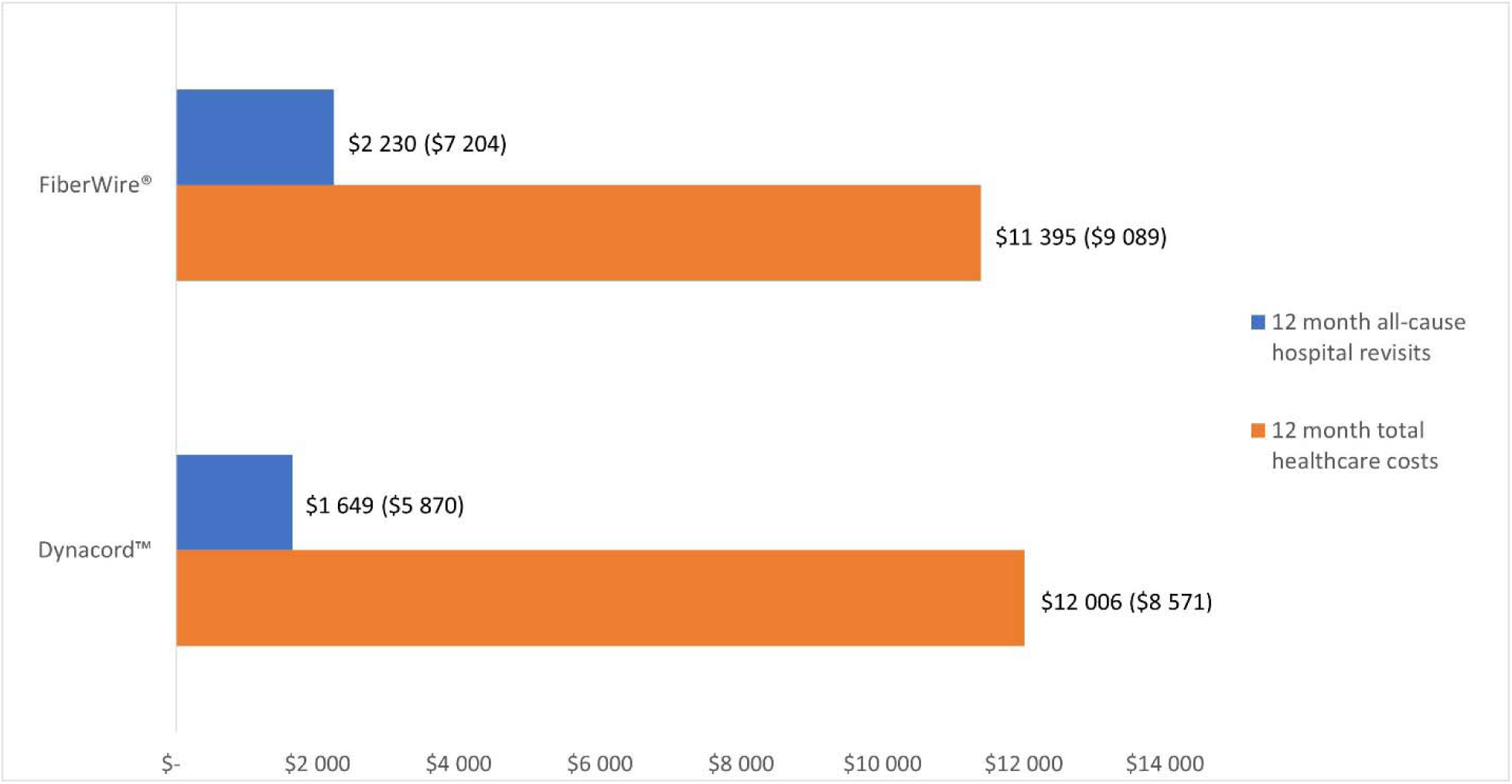
Mean (SD) cost of 12-month all-cause revisits and total costs (index costs + twelve-month revisit cost) by suture (<u>no statistical adjustment for differences in cohorts</u> <u>conducted</u>)

## Discussion

There have been significant advances in the understanding of rotator cuff disease and its management in recent years.^16^ The current study evaluated real-world patient characteristics and clinically and economically meaningful outcomes of two types of orthopedic sutures for rotator cuff repair in a hospital-based setting of care. To our knowledge, this is the first real-world data study that evaluated postoperative clinical outcomes, healthcare resource utilization, and costs associated with orthopedic sutures in a hospital setting.

Using PHD data from 1 074 patients who received Dynacord™ and 19 899 patients who received FiberWire^®^, the one-year re-repair rates in the hospital setting were 1.2% and 1.8%, respectively. A recent real-world study in a healthcare claims database by Sequeria et al. 2023 reported one-year rotator cuff re-repair rates of 4.6%, 5.2%, 8.0% among a large cohort of Medicare, Commercial, and Medicaid patients (representing ∼550 000 patients) that had open or arthroscopic rotator cuff repair.^20^ In a randomized controlled trial (RCT) of single (48 patients) versus double row (42 patients) arthroscopic rotator cuff repair, re-repair surgery was not observed in patients until two years post the first repair.^21^ Many smaller prospective and retrospective studies have measured long-term (+10 years) re-repairs in cohorts of 100 patients or less receiving a mix of open/arthroscopic rotator cuff approaches that have reported re-repair rates ranging from 3.8% to 15.4%.^22^ Unlike past research, this current study presents estimates of twelve-month re-repair occurrence expected in the inpatient and outpatient hospital setting; an estimate that has not been widely studied, but important to understanding the healthcare utilization of rotator cuff repair.

The overall incidence of twelve-month all-cause hospital revisits was 31.4% and 42.4%, Dynacord^TM^ and FiberWire^®^, respectively. Of these returns, the occurrence of patients returning for shoulder-related revisits, which may not necessarily be complications but additional returns by the patient for shoulder-related diagnoses and procedures, was 12.6% and 14.0%, Dynacord™ and FiberWire^®^, respectively. Among the shoulder-related revisits, 1.2% (Dynacord™) and 1.8% (FiberWire^®^) were for rotator cuff re-repairs, and per suture group, hospital-based complications evaluated in this study occurred in ≤ 1% of patients, respectively. Because of the data source used in this study, a hospital-based source, complications occurrence may be underestimated.

Two prior studies that evaluated one-year complications in arthroscopic and open procedure patients using retrospective studies found the overall one-year complication rates to be about 20% and 10%, arthroscopic and open, respectively.^23^ Events observed were failure to heal, stiffness, infection, nerve injury, reflex sympathetic dystrophy, deep venous thrombosis, and death. Individually, past studies of various study designs, settings of care and follow-up times have reported the occurrence of stiffness to range from 3% to 25%, superficial infection from <0.01% to 1%, deep infection from 0.03% to 3.4%, and mechanical complications such as anchor pullout from 0.01% to 3.1%.^24^

Currently, to our knowledge, published studies have not reported on healthcare utilization and costs associated with sutures for rotator cuff repair, only on the cost/utilization of the procedure itself. The real-world study performed by Sequeira et al. reported the one-year healthcare costs after rotator cuff repair from the payer perspective which were $10 843; $11 299; and $15 909 for Medicare, Commercial and Medicaid, respectively.^20^ In contrast, this current study shows the average healthcare costs from the hospital perspective in total and postoperatively (total mean hospital costs per suture were ∼$12 000 and costs postoperatively were ∼$2 000) for individual sutures.

The main strength of this study is the utilization of real-world clinically and economically meaningful data for a large population of patients treated in a hospital setting. Observational studies leverage data originating from clinical practice and better reflect real-world conditions compared to randomized controlled trials (RCTs). Although RCTs have a high degree of internal validity, they have reduced external validity as the study populations, protocols, and circumstances may not be relevant to the ‘real-world’ or to diverse populations.^25, 26^ Real-world evidence assists consumers, clinicians, purchasers, and policymakers in making informed individual- and population-level decisions.^25, 26^

A limitation of the current study is that it only captured outcomes, resource utilization, and cost in the PHD for the index hospitalization and for up to twelve-month post-rotator cuff repair in hospital settings where the patient had their index surgery. The resource use and costs incurred by patients outside of the hospital setting are not known as the PHD only contains hospitalization information, thus causing a potential underestimation of events and costs that can occur more often in nonhospital settings of care. Also, resource use and costs beyond twelve-month post-rotator cuff surgery were not evaluated, which could give important information for outcomes and resource utilization over longer periods of time. Continued research with longer-term follow-up is needed to confirm the overall clinical and economic impact of orthopedic sutures.

Another limitation of this study is that it used billing data which are not collected specifically for research purposes and which lack information (i.e., clinical and socioeconomic variables) that limit data that can be collected about the rotator cuff tear, surgery technique and the outcomes, such as rotator cuff tear type and size, single or double row technique and specific failure modes of surgery. Also, billing data may have clerical inaccuracies, recording bias secondary to financial incentives, and temporal changes in billing codes.^27, 28^ This study is also limited in that the findings from the PHD may not be generalizable to all patients having rotator cuff repair in hospital settings, particularly those in other countries.

Lastly, the study design did not include steps to provide estimates by potential confounding factors that may affect the outcome evaluated in this study such as surgeon experience, surgical technique, and rotator cuff characteristics.

## Conclusions

Despite the study limitations, the findings contribute evidence on the hospital-related real-world clinical and economic outcomes of two types of orthopedic sutures in rotator cuff repair, an area of research where evidence is limited. Incidence of re-repair in the hospital setting was 1.2% and 1.8%, Dynacord™ and FiberWire^®^, respectively. Complications, device removals, and manipulation of stiffness were uncommon. Post-operative shoulder-related visits in the hospital were 12.6% and 14.0%, Dynacord™ and FiberWire^®^, respectively. Continued research with longer-term follow-up and in non-hospital settings that accounts for baseline covariate differences in patient cohorts is needed to confirm the overall clinical and economic impact of orthopedic sutures.

## Data Availability

All data produced in the present study are available upon reasonable request to the authors

## Acknowledgements

The authors would like to thankfully acknowledge the extensive medical writing support by Natalie C. Edwards of Health Services Consulting Corporation and the extensive the computer programming support from Sauvik De and Mukesh Sharma of Cognizant LLC.,

