## Supplemental File for "Description of Patient Characteristics and Clinical and Economic Outcomes of Orthopedic Sutures Used for Rotator Cuff Repair"

### Supplemental File. CPT and ICD-10 Codes Used for Patient Identification, Patient Characteristics, Shoulder-related Revisits, Re-repairs, and Complications

Patient Identification % = means any subsequent digit

**Rotator cuff repair procedure**

| **Code type** | **Code** | **Description** |
| --- | --- | --- |
| ICD-10-PCS | 0LQ10ZZ | Repair Right Shoulder Tendon, Open Approach |
| ICD-10-PCS | 0LQ13ZZ | Repair Right Shoulder Tendon, Percutaneous Approach |
| ICD-10-PCS | 0LQ14ZZ | Repair Right Shoulder Tendon, Percutaneous Endoscopic Approach |
| ICD-10-PCS | 0LQ20ZZ | Repair Left Shoulder Tendon, Open Approach |
| ICD-10-PCS | 0LQ23ZZ | Repair Left Shoulder Tendon, Percutaneous Approach |
| ICD-10-PCS | 0LQ24ZZ | Repair Left Shoulder Tendon, Percutaneous Endoscopic Approach |
| CPT-4 | 29827 | Arthroscopy, shoulder, surgical; with rotator cuff repair |
| CPT-4 | 23410 | open acute rotator cuff repair |
| CPT-4 | 23412 | open chronic rotator cuff repair |
| ICD-10-PCS | 0LM10ZZ | Reattachment of Right Shoulder Tendon, Open Approach |
| ICD-10-PCS | 0LM14ZZ | Reattachment of Right Shoulder Tendon, Percutaneous Endoscopic Approach |
| ICD-10-PCS | 0LM20ZZ | Reattachment of Left Shoulder Tendon, Open Approach |
| ICD-10-PCS | 0LM24ZZ | Reattachment of Left Shoulder Tendon, Percutaneous Endoscopic Approach |
| **Rotator cuff diagnosis** | |  |
| **Code type** | **Code** | **Code description** |
| ICD-10-CM | M75.120 | Complete rotator cuff tear or rupture of unspecified shoulder, not specified as traumatic |
| ICD-10-CM | M75.121 | Complete rotator cuff tear or rupture of right shoulder, not specified as traumatic |
| ICD-10-CM | M75.122 | Complete rotator cuff tear or rupture of left shoulder, not specified as traumatic |
| ICD-10-CM | M75.100 | Unspecified rotator cuff tear or rupture of unspecified shoulder, not specified as traumatic |
| ICD-10-CM | M75.101 | Unspecified rotator cuff tear or rupture of right shoulder, not specified as traumatic |
| ICD-10-CM | M75.102 | Unspecified rotator cuff tear or rupture of left shoulder, not specified as traumatic |
| ICD-10-CM | M75.110 | Incomplete rotator cuff tear or rupture of unspecified shoulder, not specified as traumatic |
| ICD-10-CM | M75.111 | Incomplete rotator cuff tear or rupture of right shoulder, not specified as traumatic |
| ICD-10-CM | M75.112 | Incomplete rotator cuff tear or rupture of left shoulder, not specified as traumatic |
| ICD-10-CM | S46.001% | Unspecified injury of muscle(s) and tendon(s) of the rotator cuff of right shoulder |
| ICD-10-CM | S46.002% | Unspecified injury of muscle(s) and tendon(s) of the rotator cuff of left shoulder |
| ICD-10-CM | S46.009% | Unspecified injury of muscle(s) and tendon(s) of the rotator cuff of unspecified shoulder |
| ICD-10-CM | S46.011% | Strain of muscle(s) and tendon(s) of the rotator cuff of right shoulder |
| ICD-10-CM | S46.012% | Strain of muscle(s) and tendon(s) of the rotator cuff of left shoulder |
| ICD-10-CM | S46.019% | Strain of muscle(s) and tendon(s) of the rotator cuff of unspecified shoulder |
| ICD-10-CM | S46.021% | Laceration of muscle(s) and tendon(s) of the rotator cuff of right shoulder |
| ICD-10-CM | S46.022% | Laceration of muscle(s) and tendon(s) of the rotator cuff of left shoulder |
| ICD-10-CM | S46.029% | Laceration of muscle(s) and tendon(s) of the rotator cuff of unspecified shoulder |
| ICD-10-CM | S46.091% | Other injury of muscle(s) and tendon(s) of the rotator cuff of right shoulder |
| ICD-10-CM | S46.092% | Other injury of muscle(s) and tendon(s) of the rotator cuff of left shoulder |
| ICD-10-CM | S46.099% | Other injury of muscle(s) and tendon(s) of the rotator cuff of unspecified shoulder |

| **All codes below are ICD-10, unless specified as otherwise.**  **Diagnosis of Rotator cuff revision procedure** | |
| --- | --- |
| **Code** | **Description** |
| T84.018A | Broken internal joint prosthesis, other site, initial encounter |
| T84.019A | Broken internal joint prosthesis, unspecified site, initial encounter |
| T84.029A | Dislocation of unspecified internal joint prosthesis, initial encounter |
| T84.038A | Mechanical loosening of other internal prosthetic joint, initial encounter |
| T84.039A | Mechanical loosening of unspecified internal prosthetic joint, initial encounter |
| T84.058A | Periprosthetic osteolysis of other internal prosthetic joint, initial encounter |
| T84.098A | Other mechanical complication of other internal joint prosthesis, initial encounter |
| T84.099A | Other mechanical complication of unspecified internal joint prosthesis, initial encounter |
| T84.498A | Other mechanical complication of other internal orthopedic devices, implants and grafts, initial encounter |
| T84.81XA | Embolism due to internal orthopedic prosthetic devices, implants and grafts, initial encounter |
| T84.82XA | Fibrosis due to internal orthopedic prosthetic devices, implants and grafts, initial encounter |
| T84.83XA | Hemorrhage due to internal orthopedic prosthetic devices, implants and grafts, initial encounter |
| T84.84XA | Pain due to internal orthopedic prosthetic devices, implants and grafts, initial encounter |
| T84.85XA | Stenosis due to internal orthopedic prosthetic devices, implants and grafts, initial encounter |
| T84.86XA | Thrombosis due to internal orthopedic prosthetic devices, implants and grafts, initial encounter |
| T84.89XA | Other specified complication of internal orthopedic prosthetic devices, implants and grafts, initial encounter |
| T84.9XXA | Unspecified complication of internal orthopedic prosthetic device, implant and graft, initial encounter |
| T85.818A | Embolism due to other internal prosthetic devices, implants and grafts, initial encounter |
| T85.828A | Fibrosis due to other internal prosthetic devices, implants and grafts, initial encounter |
| T85.838A | Hemorrhage due to other internal prosthetic devices, implants and grafts, initial encounter |
| T85.848A | Pain due to other internal prosthetic devices, implants and grafts, initial encounter |
| T85.858A | Stenosis due to other internal prosthetic devices, implants and grafts, initial encounter |
| T85.868A | Thrombosis due to other internal prosthetic devices, implants and grafts, initial encounter |
| T85.898A | Other specified complication of other internal prosthetic devices, implants and grafts, initial encounter |
| T85.9XXA | Unspecified complication of internal prosthetic device, implant and graft, initial encounter |
| T84.50XA | Infection and inflammatory reaction due to unspecified internal joint prosthesis, initial encounter |
| T84.60XA | Infection and inflammatory reaction due to internal fixation device of unspecified site, initial encounter |
| T84.7XXA | Infection and inflammatory reaction due to other internal orthopedic prosthetic devices, implants and grafts, initial encounter |
| T85.79XA | Infection and inflammatory reaction due to other internal prosthetic devices, implants and grafts, initial encounter |
| **infection** |  |
| **Code** | **Description** |
| M00.011 | Staphylococcal arthritis, right shoulder |
| M00.012 | Staphylococcal arthritis, left shoulder |
| M00.019 | Staphylococcal arthritis, unspecified shoulder |
| M00.021 | Staphylococcal arthritis, right elbow |
| M00.022 | Staphylococcal arthritis, left elbow |
| M00.029 | Staphylococcal arthritis, unspecified elbow |
| M00.121 | Pneumococcal arthritis, right elbow |
| M00.122 | Pneumococcal arthritis, left elbow |
| M00.129 | Pneumococcal arthritis, unspecified elbow |
| M00.211 | Other streptococcal arthritis, right shoulder |
| M00.212 | Other streptococcal arthritis, left shoulder |
| M00.219 | Other streptococcal arthritis, unspecified shoulder |
| M00.221 | Other streptococcal arthritis, right elbow |
| M00.222 | Other streptococcal arthritis, left elbow |
| M00.229 | Other streptococcal arthritis, unspecified elbow |
| M00.811 | Arthritis due to other bacteria, right shoulder |
| M00.812 | Arthritis due to other bacteria, left shoulder |
| M00.819 | Arthritis due to other bacteria, unspecified shoulder |
| M00.821 | Arthritis due to other bacteria, right elbow |
| M00.822 | Arthritis due to other bacteria, left elbow |
| M00.829 | Arthritis due to other bacteria, unspecified elbow |
| M01.X11 | Direct infection of right shoulder in infectious and parasitic diseases classified elsewhere |
| M01.X12 | Direct infection of left shoulder in infectious and parasitic diseases classified elsewhere |
| M01.X19 | Direct infection of unspecified shoulder in infectious and parasitic diseases classified elsewhere |
| M01.X21 | Direct infection of right elbow in infectious and parasitic diseases classified elsewhere |
| M01.X22 | Direct infection of left elbow in infectious and parasitic diseases classified elsewhere |
| M01.X29 | Direct infection of unspecified elbow in infectious and parasitic diseases classified elsewhere |
| M86.119 | Other acute osteomyelitis, unspecified shoulder |
| M86.129 | Other acute osteomyelitis, unspecified humerus |
| M86.219 | Subacute osteomyelitis, unspecified shoulder |
| M86.229 | Subacute osteomyelitis, unspecified humerus |
| M86.611 | Other chronic osteomyelitis, right shoulder |
| M86.612 | Other chronic osteomyelitis, left shoulder |
| M86.619 | Other chronic osteomyelitis, unspecified shoulder |
| M86.621 | Other chronic osteomyelitis, right humerus |
| M86.622 | Other chronic osteomyelitis, left humerus |
| M86.629 | Other chronic osteomyelitis, unspecified humerus |
| M86.8X1 | Other osteomyelitis, shoulder |
| M86.8X2 | Other osteomyelitis, upper arm |
| M86.9 | Osteomyelitis, unspecified |
| M90.811 | Osteopathy in diseases classified elsewhere, right shoulder |
| M90.812 | Osteopathy in diseases classified elsewhere, left shoulder |
| M90.819 | Osteopathy in diseases classified elsewhere, unspecified shoulder |
| M90.821 | Osteopathy in diseases classified elsewhere, right upper arm |
| M90.822 | Osteopathy in diseases classified elsewhere, left upper arm |
| M90.829 | Osteopathy in diseases classified elsewhere, unspecified upper arm |
| T84.50XA | Infection and inflammatory reaction due to unspecified internal joint prosthesis, initial encounter |
| T84.60XA | Infection and inflammatory reaction due to internal fixation device of unspecified site, initial encounter |
| T84.7XXA | Infection and inflammatory reaction due to other internal orthopedic prosthetic devices, implants and grafts, initial encounter |
| T85.79XA | Infection and inflammatory reaction due to other internal prosthetic devices, implants and grafts, initial encounter |
| **shoulder fracture** | |
| **Code** | **Description** |
| S42.0% | Fracture of clavicle |
| S42.1% | Fracture of scapula |
| S42.2% | Fracture of upper end of humerus |
| S42.9% | Fracture of shoulder girdle, part unspecified |
| **fracture of any anatomy** | |
| **Code** | **Description** |
| S02.% | Fracture of skull and facial bones |
| S12.% | Fracture of cervical vertebra and other parts of neck |
| S22.% | Fracture of rib(s), sternum and thoracic spine |
| S32.% | Fracture of lumbar spine and pelvis |
| S42.% | Fracture of shoulder region |
| S52.% | Fracture of forearm |
| S62.% | Fracture at wrist and hand level |
| S72.% | Fracture of head and neck of femur |
| S82.% | Fracture of lower leg, including ankle |
| S92.% | Fracture of foot and toe, except ankle |
| *800.% to 829.% | Fracture |
| *these represent ICD-9 codes. | |

**Clinical status (degenerative or trauma)**

| **Code** | **Code description** | **trauma** |
| --- | --- | --- |
| M75.120 | Complete rotator cuff tear or rupture of unspecified shoulder, not specified as traumatic | degenerative |
| M75.121 | Complete rotator cuff tear or rupture of right shoulder, not specified as traumatic | degenerative |
| M75.122 | Complete rotator cuff tear or rupture of left shoulder, not specified as traumatic | degenerative |
| M75.100 | Unspecified rotator cuff tear or rupture of unspecified shoulder, not specified as traumatic | degenerative |
| M75.101 | Unspecified rotator cuff tear or rupture of right shoulder, not specified as traumatic | degenerative |
| M75.102 | Unspecified rotator cuff tear or rupture of left shoulder, not specified as traumatic | degenerative |
| M75.110 | Incomplete rotator cuff tear or rupture of unspecified shoulder, not specified as traumatic | degenerative |
| M75.111 | Incomplete rotator cuff tear or rupture of right shoulder, not specified as traumatic | degenerative |
| M75.112 | Incomplete rotator cuff tear or rupture of left shoulder, not specified as traumatic | degenerative |
| S46.001% | Unspecified injury of muscle(s) and tendon(s) of the rotator cuff of right shoulder | Trauma |
| S46.002% | Unspecified injury of muscle(s) and tendon(s) of the rotator cuff of left shoulder | Trauma |
| S46.009% | Unspecified injury of muscle(s) and tendon(s) of the rotator cuff of unspecified shoulder | Trauma |
| S46.011% | Strain of muscle(s) and tendon(s) of the rotator cuff of right shoulder | Trauma |
| S46.012% | Strain of muscle(s) and tendon(s) of the rotator cuff of left shoulder | Trauma |
| S46.019% | Strain of muscle(s) and tendon(s) of the rotator cuff of unspecified shoulder | Trauma |
| S46.021% | Laceration of muscle(s) and tendon(s) of the rotator cuff of right shoulder | Trauma |
| S46.022% | Laceration of muscle(s) and tendon(s) of the rotator cuff of left shoulder | Trauma |
| S46.029% | Laceration of muscle(s) and tendon(s) of the rotator cuff of unspecified shoulder | Trauma |
| S46.091% | Other injury of muscle(s) and tendon(s) of the rotator cuff of right shoulder | Trauma |
| S46.092% | Other injury of muscle(s) and tendon(s) of the rotator cuff of left shoulder | Trauma |
| S46.099% | Other injury of muscle(s) and tendon(s) of the rotator cuff of unspecified shoulder | Trauma |

Tobacco use

| **Code** | **Description** |
| --- | --- |
| 99406 | Smoking and tobacco use cessation counseling visit is greater than three minutes, but not more than 10 minutes |
| 99407 | Smoking and tobacco use cessation counseling visit is greater than 10 minutes |
| S9075 | Smoking cessation treatment |
| S9453 | Smoking cessation classes |
| G0436 | Smoking and tobacco use cessation counseling visit greater than three minutes, but not more than 10 minutes. |
| G0437 | Smoking and tobacco use cessation counseling visit is greater than 10 minutes. |
| F17.2XX | Nicotine dependence |
| Z87.891 | Personal history of nicotine dependence |
| Z72.0XX | Tobacco use |
| Z71.6XX | Tobacco abuse counseling |

**Approach and Laterality**

***note: for laterality variable assign right, left, unknown using ICD-10 procedure, CPT-4 or ICD-10 diagnosis Codes**

**ICD-10 procedure codes and CPT-4**

| **Code** | **Description** | **approach** | **Laterality*** |
| --- | --- | --- | --- |
| 0LQ10ZZ | Repair Right Shoulder Tendon, Open Approach | open | Right |
| 0LQ13ZZ | Repair Right Shoulder Tendon, Percutaneous Approach | arthroscopic | Right |
| 0LQ14ZZ | Repair Right Shoulder Tendon, Percutaneous Endoscopic Approach | arthroscopic | Right |
| 0LQ20ZZ | Repair Left Shoulder Tendon, Open Approach | open | Left |
| 0LQ23ZZ | Repair Left Shoulder Tendon, Percutaneous Approach | arthroscopic | Left |
| 0LQ24ZZ | Repair Left Shoulder Tendon, Percutaneous Endoscopic Approach | arthroscopic | Left |
| 29827 | Arthroscopy, shoulder, surgical; with rotator cuff repair | arthroscopic | depends on modifer or diagnostic codes or unknown |
| 23410 | open acute rotator cuff repair | open | depends on modifer or diagnostic codes or unknown |
| 23412 | open chronic rotator cuff repair | open | depends on modifer or diagnostic codes or unknown |
| 0LM10ZZ | Reattachment of Right Shoulder Tendon, Open Approach | open | Right |
| 0LM14ZZ | Reattachment of Right Shoulder Tendon, Percutaneous Endoscopic Approach | arthroscopic | Right |
| 0LM20ZZ | Reattachment of Left Shoulder Tendon, Open Approach | open | Left |
| 0LM24ZZ | Reattachment of Left Shoulder Tendon, Percutaneous Endoscopic Approach | arthroscopic | Left |

**Diagnosis codes**

| **Code** | **Code description** | **Laterality*** |
| --- | --- | --- |
| M75.120 | Complete rotator cuff tear or rupture of unspecified shoulder, not specified as traumatic | unknown |
| M75.121 | Complete rotator cuff tear or rupture of right shoulder, not specified as traumatic | right |
| M75.122 | Complete rotator cuff tear or rupture of left shoulder, not specified as traumatic | left |
| M75.100 | Unspecified rotator cuff tear or rupture of unspecified shoulder, not specified as traumatic | unknown |
| M75.101 | Unspecified rotator cuff tear or rupture of right shoulder, not specified as traumatic | right |
| M75.102 | Unspecified rotator cuff tear or rupture of left shoulder, not specified as traumatic | left |
| M75.110 | Incomplete rotator cuff tear or rupture of unspecified shoulder, not specified as traumatic | unknown |
| M75.111 | Incomplete rotator cuff tear or rupture of right shoulder, not specified as traumatic | right |
| M75.112 | Incomplete rotator cuff tear or rupture of left shoulder, not specified as traumatic | left |
| S46.001% | Unspecified injury of muscle(s) and tendon(s) of the rotator cuff of right shoulder | right |
| S46.002% | Unspecified injury of muscle(s) and tendon(s) of the rotator cuff of left shoulder | left |
| S46.009% | Unspecified injury of muscle(s) and tendon(s) of the rotator cuff of unspecified shoulder | unknown |
| S46.011% | Strain of muscle(s) and tendon(s) of the rotator cuff of right shoulder | right |
| S46.012% | Strain of muscle(s) and tendon(s) of the rotator cuff of left shoulder | left |
| S46.019% | Strain of muscle(s) and tendon(s) of the rotator cuff of unspecified shoulder | unknown |
| S46.021% | Laceration of muscle(s) and tendon(s) of the rotator cuff of right shoulder | right |
| S46.022% | Laceration of muscle(s) and tendon(s) of the rotator cuff of left shoulder | left |
| S46.029% | Laceration of muscle(s) and tendon(s) of the rotator cuff of unspecified shoulder | unknown |
| S46.091% | Other injury of muscle(s) and tendon(s) of the rotator cuff of right shoulder | right |
| S46.092% | Other injury of muscle(s) and tendon(s) of the rotator cuff of left shoulder | left |
| S46.099% | Other injury of muscle(s) and tendon(s) of the rotator cuff of unspecified shoulder | unknown |

**Rotator cuff re-repair**

| **Code type** | **Code** | **Description** |
| --- | --- | --- |
| ICD-10-PCS | 0LQ10ZZ | Repair Right Shoulder Tendon, Open Approach |
| ICD-10-PCS | 0LQ13ZZ | Repair Right Shoulder Tendon, Percutaneous Approach |
| ICD-10-PCS | 0LQ14ZZ | Repair Right Shoulder Tendon, Percutaneous Endoscopic Approach |
| ICD-10-PCS | 0LQ20ZZ | Repair Left Shoulder Tendon, Open Approach |
| ICD-10-PCS | 0LQ23ZZ | Repair Left Shoulder Tendon, Percutaneous Approach |
| ICD-10-PCS | 0LQ24ZZ | Repair Left Shoulder Tendon, Percutaneous Endoscopic Approach |
| CPT-4 | 29827 | Arthroscopy, shoulder, surgical; with rotator cuff repair |
| CPT-4 | 23410 | Repair of ruptured musculotendinous cuff (eg, rotator cuff) open; acute |
| CPT-4 | 23412 | Repair of ruptured musculotendinous cuff (eg, rotator cuff) open; chronic |
| CPT-4 | 23420 | Reconstruction of complete shoulder (rotator) cuff avulsion chronic (includes acromioplasty) |
| ICD-10-PCS | 0LM10ZZ | Reattachment of Right Shoulder Tendon, Open Approach |
| ICD-10-PCS | 0LM14ZZ | Reattachment of Right Shoulder Tendon, Percutaneous Endoscopic Approach |
| ICD-10-PCS | 0LM20ZZ | Reattachment of Left Shoulder Tendon, Open Approach |
| ICD-10-PCS | 0LM24ZZ | Reattachment of Left Shoulder Tendon, Percutaneous Endoscopic Approach |

**Manipulation of stiffness**

| **Code type** | **Code** | **Description** |
| --- | --- | --- |
| ICD-10-PCS | 0RSEXZZ | Reposition Right Sternoclavicular Joint, External Approach |
| ICD-10-PCS | 0RSFXZZ | Reposition left Sternoclavicular Joint, External Approach |
| ICD-10-PCS | 0RSGXZZ | Reposition Right Acromioclavicular Joint, External Approach |
| ICD-10-PCS | 0RSHXZZ | Reposition Left Acromioclavicular Joint, External Approach |
| ICD-10-PCS | 0RSKXZZ | Reposition Left Shoulder Joint, External Approach |
| ICD-10-PCS | 0RSJXZZ | Reposition Right Shoulder Joint, External Approach |
| ICD-10-PCS | 0RNEXZZ | Release Right Sternoclavicular Joint, External Approach |
| ICD-10-PCS | 0RNFXZZ | Release Left Sternoclavicular Joint, External Approach |
| ICD-10-PCS | 0RNGXZZ | Release Right Acromioclavicular Joint, External Approach |
| ICD-10-PCS | 0RNHXZZ | Release Left Acromioclavicular Joint, External Approach |
| ICD-10-PCS | 0RNJXZZ | Release Right Shoulder Joint, External Approach |
| ICD-10-PCS | 0RNKXZZ | Release Left Shoulder Joint, External Approach |
| CPT-4 | 23700 | Manipulation under anesthesia, shoulder joint, including application of fixation apparatus (dislocation excluded) |

**Device removal**

| ICD-10-PCS | 0LPX0YZ | Removal of Other Device from Upper Tendon, Open Approach |
| --- | --- | --- |
| ICD-10-PCS | 0LPX3YZ | Removal of Other Device from Upper Tendon,Percutaneous Approach |
| ICD-10-PCS | 0LPX4YZ | Removal of Other Device from Upper Tendon,Percutaneous Approach Endoscopic |

**Complications**

| **Tissue cutting** | |  |
| --- | --- | --- |
| **Code type** | **Code** | **Description** |
| ICD-10-CM | T85.898A | Other specified complication of other internal prosthetic devices, implants and grafts, initial encounter |
| ICD-10-CM | T84.89XA | Other specified complication of internal orthopedic prosthetic devices, implants and grafts, initial encounter |
| ICD-10-CM | Y79.3 | Orthopedic devices associated with adverse incidents, Surgical instruments, materials and orthopedic devices (including sutures) associated with adverse incidents |
| **Foreign body reaction** | | |
| **Code type** | **Code** | **Description** |
| ICD-10-CM | T78.49XA | Other allergy, initial encounter |
| ICD-10-CM | T85.898A | Other specified complication of other internal prosthetic devices, implants and grafts, initial encounter |
| **Mechanical comps** | | |
| **Code type** | **Code** | **Description** |
| ICD-10-CM | T84.498A | Other mechanical complication of other internal orthopedic devices, implants and grafts, initial encounter |
| **Infection** | | |
| **Code type** | **Code** | **Description** |
| ICD-10-CM | M86111 | Other acute osteomyelitis right shoulder |
| ICD-10-CM | M86112 | Other acute osteomyelitis left shoulder |
| ICD-10-CM | M86119 | Other acute osteomyelitis unspecified shoulder |
| ICD-10-CM | M86121 | Other acute osteomyelitis right humerus |
| ICD-10-CM | M86122 | Other acute osteomyelitis left humerus |
| ICD-10-CM | M86129 | Other acute osteomyelitis unspecified humerus |
| ICD-10-CM | M86211 | Subacute osteomyelitis right shoulder |
| ICD-10-CM | M86212 | Subacute osteomyelitis left shoulder |
| ICD-10-CM | M86219 | Subacute osteomyelitis unspecified shoulder |
| ICD-10-CM | M86221 | Subacute osteomyelitis right humerus |
| ICD-10-CM | M86222 | Subacute osteomyelitis left humerus |
| ICD-10-CM | M86229 | Subacute osteomyelitis unspecified humerus |
| ICD-10-CM | M86611 | Other chronic osteomyelitis right shoulder |
| ICD-10-CM | M86612 | Other chronic osteomyelitis left shoulder |
| ICD-10-CM | M86619 | Other chronic osteomyelitis unspecified shoulder |
| ICD-10-CM | M86621 | Other chronic osteomyelitis right humerus |
| ICD-10-CM | M86622 | Other chronic osteomyelitis left humerus |
| ICD-10-CM | M86629 | Other chronic osteomyelitis unspecified humerus |
| ICD-10-CM | T8140XA | Infection following a procedure, unspecified, initial encounter |
| ICD-10-CM | T8141XA | Infection following a procedure, superficial incisional surgical site, initial encounter |
| ICD-10-CM | T8142XA | Infection following a procedure, deep incisional surgical site, initial encounter |
| ICD-10-CM | T8143XA | Infection following a procedure, organ and space surgical site, initial encounter |
| ICD-10-CM | T8144XA | Sepsis following a procedure, initial encounter |
| ICD-10-CM | T8149XA | Infection following a procedure, other surgical site, initial encounter |
| ICD-10-CM | T814XXA | Infection following a procedure, initial encounter |
| ICD-10-CM | T84610A | INFECTION AND INFLAMMATORY REACTION DUE TO INTERNAL FIXATION DEVICE OF RIGHT HUMERUS INITIAL ENCOUNTER |
| ICD-10-CM | T84611A | INFECTION AND INFLAMMATORY REACTION DUE TO INTERNAL FIXATION DEVICE OF LEFT HUMERUS INITIAL ENCOUNTER |
| ICD-10-CM | T84.59XA | Infection and inflammatory reaction due to other internal joint prosthesis, initial encounter |

**Shoulder-related revisits**

**Business rule: include all codes below and re-repair, manipulation of stiffness, device removal, and complication codes in tables above.**

| **Code type** | **Code** | **Description** |
| --- | --- | --- |
| ICD-10-CM | S42101A | FRACTURE OF UNSPECIFIED PART OF SCAPULA RIGHT SHOULDER INITIAL ENCOUNTER FOR CLOSED FRACTURE |
| ICD-10-CM | S42102A | FRACTURE OF UNSPECIFIED PART OF SCAPULA LEFT SHOULDER INITIAL ENCOUNTER FOR CLOSED FRACTURE |
| ICD-10-CM | S42109A | FRACTURE OF UNSPECIFIED PART OF SCAPULA UNSPECIFIED SHOULDER INITIAL ENCOUNTER FOR CLOSED FRACTURE |
| ICD-10-CM | S42111A | DISPLACED FRACTURE OF BODY OF SCAPULA RIGHT SHOULDER INITIAL ENCOUNTER FOR CLOSED FRACTURE |
| ICD-10-CM | S42112A | DISPLACED FRACTURE OF BODY OF SCAPULA LEFT SHOULDER INITIAL ENCOUNTER FOR CLOSED FRACTURE |
| ICD-10-CM | S42113A | DISPLACED FRACTURE OF BODY OF SCAPULA UNSPECIFIED SHOULDER INITIAL ENCOUNTER FOR CLOSED FRACTURE |
| ICD-10-CM | S42114A | NONDISPLACED FRACTURE OF BODY OF SCAPULA RIGHT SHOULDER INITIAL ENCOUNTER FOR CLOSED FRACTURE |
| ICD-10-CM | S42115A | NONDISPLACED FRACTURE OF BODY OF SCAPULA LEFT SHOULDER INITIAL ENCOUNTER FOR CLOSED FRACTURE |
| ICD-10-CM | S42116A | NONDISPLACED FRACTURE OF BODY OF SCAPULA UNSPECIFIED SHOULDER INITIAL ENCOUNTER FOR CLOSED FRACTURE |
| ICD-10-CM | S42141A | DISPLACED FRACTURE OF GLENOID CAVITY OF SCAPULA RIGHT SHOULDER INITIAL ENCOUNTER FOR CLOSED FRACTURE |
| ICD-10-CM | S42142A | DISPLACED FRACTURE OF GLENOID CAVITY OF SCAPULA LEFT SHOULDER INITIAL ENCOUNTER FOR CLOSED FRACTURE |
| ICD-10-CM | S42143A | DISPLACED FRACTURE OF GLENOID CAVITY OF SCAPULA UNSPECIFIED SHOULDER INITIAL ENCOUNTER FOR CLOSED FRACTURE |
| ICD-10-CM | S42144A | NONDISPLACED FRACTURE OF GLENOID CAVITY OF SCAPULA RIGHT SHOULDER INITIAL ENCOUNTER FOR CLOSED FRACTURE |
| ICD-10-CM | S42145A | NONDISPLACED FRACTURE OF GLENOID CAVITY OF SCAPULA LEFT SHOULDER INITIAL ENCOUNTER FOR CLOSED FRACTURE |
| ICD-10-CM | S42146A | NONDISPLACED FRACTURE OF GLENOID CAVITY OF SCAPULA UNSPECIFIED SHOULDER INITIAL ENCOUNTER FOR CLOSED FRACTURE |
| ICD-10-CM | S42151A | DISPLACED FRACTURE OF NECK OF SCAPULA RIGHT SHOULDER INITIAL ENCOUNTER FOR CLOSED FRACTURE |
| ICD-10-CM | S42152A | DISPLACED FRACTURE OF NECK OF SCAPULA LEFT SHOULDER INITIAL ENCOUNTER FOR CLOSED FRACTURE |
| ICD-10-CM | S42153A | DISPLACED FRACTURE OF NECK OF SCAPULA UNSPECIFIED SHOULDER INITIAL ENCOUNTER FOR CLOSED FRACTURE |
| ICD-10-CM | S42154A | NONDISPLACED FRACTURE OF NECK OF SCAPULA RIGHT SHOULDER INITIAL ENCOUNTER FOR CLOSED FRACTURE |
| ICD-10-CM | S42155A | NONDISPLACED FRACTURE OF NECK OF SCAPULA LEFT SHOULDER INITIAL ENCOUNTER FOR CLOSED FRACTURE |
| ICD-10-CM | S42156A | NONDISPLACED FRACTURE OF NECK OF SCAPULA UNSPECIFIED SHOULDER INITIAL ENCOUNTER FOR CLOSED FRACTURE |
| ICD-10-CM | S42191A | FRACTURE OF OTHER PART OF SCAPULA RIGHT SHOULDER INITIAL ENCOUNTER FOR CLOSED FRACTURE |
| ICD-10-CM | S42192A | FRACTURE OF OTHER PART OF SCAPULA LEFT SHOULDER INITIAL ENCOUNTER FOR CLOSED FRACTURE |
| ICD-10-CM | S42199A | FRACTURE OF OTHER PART OF SCAPULA UNSPECIFIED SHOULDER INITIAL ENCOUNTER FOR CLOSED FRACTURE |
| ICD-10-CM | M24311 | PATHOLOGICAL DISLOCATION OF RIGHT SHOULDER NOT ELSEWHERE CLASSIFIED |
| ICD-10-CM | M24312 | PATHOLOGICAL DISLOCATION OF LEFT SHOULDER NOT ELSEWHERE CLASSIFIED |
| ICD-10-CM | M24411 | RECURRENT DISLOCATION RIGHT SHOULDER |
| ICD-10-CM | M24412 | RECURRENT DISLOCATION LEFT SHOULDER |
| ICD-10-CM | S43% | Subluxation and dislocation of shoulder joint |
| ICD-10-CM | M01X11 | DIRECT INFECTION OF RIGHT SHOULDER IN INFECTIOUS AND PARASITIC DISEASES CLASSIFIED ELSEWHERE |
| ICD-10-CM | M01X12 | DIRECT INFECTION OF LEFT SHOULDER IN INFECTIOUS AND PARASITIC DISEASES CLASSIFIED ELSEWHERE |
| ICD-10-CM | S41001A | UNSPECIFIED OPEN WOUND OF RIGHT SHOULDER INITIAL ENCOUNTER |
| ICD-10-CM | S41002A | UNSPECIFIED OPEN WOUND OF LEFT SHOULDER INITIAL ENCOUNTER |
| ICD-10-CM | M89511 | OSTEOLYSIS RIGHT SHOULDER |
| ICD-10-CM | M89512 | OSTEOLYSIS LEFT SHOULDER |
| ICD-10-CM | M89521 | OSTEOLYSIS RIGHT Upper Arm |
| ICD-10-CM | M89522 | OSTEOLYSIS LEFT Upper Arm |
| ICD-10-CM | M9731XA | Periprosthetic fracture around internal prosthetic right shoulder joint, initial encounter |
| ICD-10-CM | M9732XA | Periprosthetic fracture around internal prosthetic left shoulder joint, initial encounter |
| ICD-10-CM | S42209A | UNSPECIFIED FRACTURE OF UPPER END OF UNSPECIFIED HUMERUS INITIAL ENCOUNTER FOR CLOSED FRACTURE |
| ICD-10-CM | S42399A | OTHER FRACTURE OF SHAFT OF UNSPECIFIED HUMERUS INITIAL ENCOUNTER FOR CLOSED FRACTURE |
| ICD-10-CM | M24411 | RECURRENT DISLOCATION RIGHT SHOULDER |
| ICD-10-CM | M24412 | RECURRENT DISLOCATION LEFT SHOULDER |
| ICD-10-CM | M24419 | RECURRENT DISLOCATION UNSPECIFIED SHOULDER |
| ICD-10-CM | T84029A | DISLOCATION OF UNSPECIFIED INTERNAL JOINT PROSTHESIS INITIAL ENCOUNTER |
| ICD-10-CM | T8459XA | INFECTION AND INFLAMMATORY REACTION DUE TO OTHER INTERNAL JOINT PROSTHESIS INITIAL ENCOUNTER |
| ICD-10-CM | T84019A | BROKEN INTERNAL JOINT PROSTHESIS UNSPECIFIED SITE INITIAL ENCOUNTER |
| ICD-10-CM | T84038A | MECHANICAL LOOSENING OF OTHER INTERNAL PROSTHETIC JOINT INITIAL ENCOUNTER |
| ICD-10-CM | T84039A | MECHANICAL LOOSENING OF UNSPECIFIED INTERNAL PROSTHETIC JOINT INITIAL ENCOUNTER |
| ICD-10-CM | M24611 | ANKYLOSIS RIGHT SHOULDER |
| ICD-10-CM | M24612 | ANKYLOSIS LEFT SHOULDER |
| ICD-10-CM | M24619 | ANKYLOSIS UNSPECIFIED SHOULDER |
| ICD-10-CM | M25611 | STIFFNESS OF RIGHT SHOULDER NOT ELSEWHERE CLASSIFIED |
| ICD-10-CM | M25612 | STIFFNESS OF LEFT SHOULDER NOT ELSEWHERE CLASSIFIED |
| ICD-10-CM | M25619 | STIFFNESS OF UNSPECIFIED SHOULDER NOT ELSEWHERE CLASSIFIED |
| ICD-10-CM | M7500 | ADHESIVE CAPSULITIS OF UNSPECIFIED SHOULDER |
| ICD-10-CM | M7501 | ADHESIVE CAPSULITIS OF RIGHT SHOULDER |
| ICD-10-CM | M7502 | ADHESIVE CAPSULITIS OF LEFT SHOULDER |
| ICD-10-CM | M75101 | UNSPECIFIED ROTATOR CUFF TEAR OR RUPTURE OF RIGHT SHOULDER NOT SPECIFIED AS TRAUMATIC |
| ICD-10-CM | M75102 | UNSPECIFIED ROTATOR CUFF TEAR OR RUPTURE OF LEFT SHOULDER NOT SPECIFIED AS TRAUMATIC |
| ICD-10-CM | M75111 | INCOMPLETE ROTATOR CUFF TEAR OR RUPTURE OF RIGHT SHOULDER NOT SPECIFIED AS TRAUMATIC |
| ICD-10-CM | M75112 | INCOMPLETE ROTATOR CUFF TEAR OR RUPTURE OF LEFT SHOULDER NOT SPECIFIED AS TRAUMATIC |
| ICD-10-CM | M75121 | COMPLETE ROTATOR CUFF TEAR OR RUPTURE OF RIGHT SHOULDER NOT SPECIFIED AS TRAUMATIC |
| ICD-10-CM | M75122 | COMPLETE ROTATOR CUFF TEAR OR RUPTURE OF LEFT SHOULDER NOT SPECIFIED AS TRAUMATIC |
| ICD-10-CM | S43421A | SPRAIN OF RIGHT ROTATOR CUFF CAPSULE INITIAL ENCOUNTER |
| ICD-10-CM | S43422A | SPRAIN OF LEFT ROTATOR CUFF CAPSULE INITIAL ENCOUNTER |
| ICD-10-CM | S46001A | UNSPECIFIED INJURY OF MUSCLE(S) AND TENDON(S) OF THE ROTATOR CUFF OF RIGHT SHOULDER INITIAL ENCOUNTER |
| ICD-10-CM | S46002A | UNSPECIFIED INJURY OF MUSCLE(S) AND TENDON(S) OF THE ROTATOR CUFF OF LEFT SHOULDER INITIAL ENCOUNTER |
| ICD-10-CM | S46011A | STRAIN OF MUSCLE(S) AND TENDON(S) OF THE ROTATOR CUFF OF RIGHT SHOULDER INITIAL ENCOUNTER |
| ICD-10-CM | S46012A | STRAIN OF MUSCLE(S) AND TENDON(S) OF THE ROTATOR CUFF OF LEFT SHOULDER INITIAL ENCOUNTER |
| ICD-10-CM | S46021A | LACERATION OF MUSCLE(S) AND TENDON(S) OF THE ROTATOR CUFF OF RIGHT SHOULDER INITIAL ENCOUNTER |
| ICD-10-CM | S46022A | LACERATION OF MUSCLE(S) AND TENDON(S) OF THE ROTATOR CUFF OF LEFT SHOULDER INITIAL ENCOUNTER |
| ICD-10-CM | S46091A | OTHER INJURY OF MUSCLE(S) AND TENDON(S) OF THE ROTATOR CUFF OF RIGHT SHOULDER INITIAL ENCOUNTER |
| ICD-10-CM | S46092A | OTHER INJURY OF MUSCLE(S) AND TENDON(S) OF THE ROTATOR CUFF OF LEFT SHOULDER INITIAL ENCOUNTER |
| ICD-10-CM | M75100 | UNSPECIFIED ROTATOR CUFF TEAR OR RUPTURE OF UNSPECIFIED SHOULDER NOT SPECIFIED AS TRAUMATIC |
| ICD-10-CM | M7550 | BURSITIS OF UNSPECIFIED SHOULDER |
| ICD-10-CM | M7551 | BURSITIS OF RIGHT SHOULDER |
| ICD-10-CM | M7552 | BURSITIS OF LEFT SHOULDER |
| ICD-10-CM | S46019A | STRAIN OF MUSCLE(S) AND TENDON(S) OF THE ROTATOR CUFF OF UNSPECIFIED SHOULDER INITIAL ENCOUNTER |
| ICD-10-CM | S46021A | LACERATION OF MUSCLE(S) AND TENDON(S) OF THE ROTATOR CUFF OF RIGHT SHOULDER INITIAL ENCOUNTER |
| ICD-10-CM | S46022A | LACERATION OF MUSCLE(S) AND TENDON(S) OF THE ROTATOR CUFF OF LEFT SHOULDER INITIAL ENCOUNTER |
| ICD-10-CM | S46029A | LACERATION OF MUSCLE(S) AND TENDON(S) OF THE ROTATOR CUFF OF UNSPECIFIED SHOULDER INITIAL ENCOUNTER |
| ICD-10-CM | S46091A | OTHER INJURY OF MUSCLE(S) AND TENDON(S) OF THE ROTATOR CUFF OF RIGHT SHOULDER INITIAL ENCOUNTER |
| ICD-10-CM | S46092A | OTHER INJURY OF MUSCLE(S) AND TENDON(S) OF THE ROTATOR CUFF OF LEFT SHOULDER INITIAL ENCOUNTER |
| ICD-10-CM | S46099A | OTHER INJURY OF MUSCLE(S) AND TENDON(S) OF THE ROTATOR CUFF OF UNSPECIFIED SHOULDER INITIAL ENCOUNTER |
| ICD-10-CM | S46001A | UNSPECIFIED INJURY OF MUSCLE(S) AND TENDON(S) OF THE ROTATOR CUFF OF RIGHT SHOULDER INITIAL ENCOUNTER |
| ICD-10-CM | S46002A | UNSPECIFIED INJURY OF MUSCLE(S) AND TENDON(S) OF THE ROTATOR CUFF OF LEFT SHOULDER INITIAL ENCOUNTER |
| ICD-10-CM | S46009A | UNSPECIFIED INJURY OF MUSCLE(S) AND TENDON(S) OF THE ROTATOR CUFF OF UNSPECIFIED SHOULDER INITIAL ENCOUNTER |
| ICD-10-CM | S46011A | STRAIN OF MUSCLE(S) AND TENDON(S) OF THE ROTATOR CUFF OF RIGHT SHOULDER INITIAL ENCOUNTER |
| ICD-10-CM | S46012A | STRAIN OF MUSCLE(S) AND TENDON(S) OF THE ROTATOR CUFF OF LEFT SHOULDER INITIAL ENCOUNTER |
| ICD-10-CM | S46019A | STRAIN OF MUSCLE(S) AND TENDON(S) OF THE ROTATOR CUFF OF UNSPECIFIED SHOULDER INITIAL ENCOUNTER |
| ICD-10-CM | S46021A | LACERATION OF MUSCLE(S) AND TENDON(S) OF THE ROTATOR CUFF OF RIGHT SHOULDER INITIAL ENCOUNTER |
| ICD-10-CM | S46022A | LACERATION OF MUSCLE(S) AND TENDON(S) OF THE ROTATOR CUFF OF LEFT SHOULDER INITIAL ENCOUNTER |
| ICD-10-CM | S46029A | LACERATION OF MUSCLE(S) AND TENDON(S) OF THE ROTATOR CUFF OF UNSPECIFIED SHOULDER INITIAL ENCOUNTER |
| ICD-10-CM | S46091A | OTHER INJURY OF MUSCLE(S) AND TENDON(S) OF THE ROTATOR CUFF OF RIGHT SHOULDER INITIAL ENCOUNTER |
| ICD-10-CM | S46092A | OTHER INJURY OF MUSCLE(S) AND TENDON(S) OF THE ROTATOR CUFF OF LEFT SHOULDER INITIAL ENCOUNTER |
| ICD-10-CM | S46099A | OTHER INJURY OF MUSCLE(S) AND TENDON(S) OF THE ROTATOR CUFF OF UNSPECIFIED SHOULDER INITIAL ENCOUNTER |
| ICD-10-CM | M7580 | OTHER SHOULDER LESIONS UNSPECIFIED SHOULDER |
| ICD-10-CM | M7581 | OTHER SHOULDER LESIONS RIGHT SHOULDER |
| ICD-10-CM | M7582 | OTHER SHOULDER LESIONS LEFT SHOULDER |
| CPT-4 | 23000 to 24079 | Shoulder procedures |
| CPT-4 | 29805 to 29828 | Shoulder arthroscopy |
| ICD-10-PCS | 0P55% | Destruction / Scapula, Right |
| ICD-10-PCS | 0P56% | Destruction / Scapula, Left |
| ICD-10-PCS | 0P55% | Destruction / Scapula, Right |
| ICD-10-PCS | 0P56% | Destruction / Scapula, Left |
| ICD-10-PCS | 0P57% | Destruction / Glenoid Cavity, Right |
| ICD-10-PCS | 0P58% | Destruction / Glenoid Cavity, Left |
| ICD-10-PCS | 0P59% | Destruction / Clavicle, Right |
| ICD-10-PCS | 0P5B% | Destruction / Clavicle, Left |
| ICD-10-PCS | 0P5C% | Destruction / Humeral Head, Right |
| ICD-10-PCS | 0P5D% | Destruction / Humeral Head, Left |
| ICD-10-PCS | 0R5E% | Destruction / Sternoclavicular Joint, Right |
| ICD-10-PCS | 0R5F% | Destruction / Sternoclavicular Joint, Left |
| ICD-10-PCS | 0R5G% | Destruction / Acromioclavicular Joint, Right |
| ICD-10-PCS | 0R5H% | Destruction / Acromioclavicular Joint, Left |
| ICD-10-PCS | 0R5J% | Destruction / Shoulder Joint, Right |
| ICD-10-PCS | 0R5K% | Destruction / Shoulder Joint, Left |
| ICD-10-PCS | 0P85% | Division / Scapula, Right |
| ICD-10-PCS | 0P86% | Division / Scapula, Left |
| ICD-10-PCS | 0P87% | Division / Glenoid Cavity, Right |
| ICD-10-PCS | 0P88% | Division / Glenoid Cavity, Left |
| ICD-10-PCS | 0P89% | Division / Clavicle, Right |
| ICD-10-PCS | 0P8B% | Division / Clavicle, Left |
| ICD-10-PCS | 0P8C% | Division / Humeral Head, Right |
| ICD-10-PCS | 0P8D% | Division / Humeral Head, Left |
| ICD-10-PCS | 0P8F% | Division / Humeral Shaft, Right |
| ICD-10-PCS | 0P8G% | Division / Humeral Shaft, Left |
| ICD-10-PCS | 0R9E% | Drainage / Sternoclavicular Joint, Right |
| ICD-10-PCS | 0R9F% | Drainage / Sternoclavicular Joint, Left |
| ICD-10-PCS | 0R9G% | Drainage / Acromioclavicular Joint, Right |
| ICD-10-PCS | 0R9H% | Drainage / Acromioclavicular Joint, Left |
| ICD-10-PCS | 0R9J% | Drainage / Shoulder Joint, Right |
| ICD-10-PCS | 0R9K% | Drainage / Shoulder Joint, Left |
| ICD-10-PCS | 0P95% | Drainage / Scapula, Right |
| ICD-10-PCS | 0P96% | Drainage / Scapula, Left |
| ICD-10-PCS | 0P97% | Drainage / Glenoid Cavity, Right |
| ICD-10-PCS | 0P98% | Drainage / Glenoid Cavity, Left |
| ICD-10-PCS | 0P99% | Drainage / Clavicle, Right |
| ICD-10-PCS | 0P9B% | Drainage / Clavicle, Left |
| ICD-10-PCS | 0P9C% | Drainage / Humeral Head, Right |
| ICD-10-PCS | 0P9D% | Drainage / Humeral Head, Left |
| ICD-10-PCS | 0P9F% | Drainage / Humeral Shaft, Right |
| ICD-10-PCS | 0P9G% | Drainage / Humeral Shaft, Left |
| ICD-10-PCS | 0PB5% | Excision / Scapula, Right |
| ICD-10-PCS | 0PB6% | Excision / Scapula, Left |
| ICD-10-PCS | 0PB7% | Excision / Glenoid Cavity, Right |
| ICD-10-PCS | 0PB8% | Excision / Glenoid Cavity, Left |
| ICD-10-PCS | 0PB9% | Excision / Clavicle, Right |
| ICD-10-PCS | 0PBB% | Excision / Clavicle, Left |
| ICD-10-PCS | 0PBC% | Excision / Humeral Head, Right |
| ICD-10-PCS | 0PBD% | Excision / Humeral Head, Left |
| ICD-10-PCS | 0PBF% | Excision / Humeral Shaft, Right |
| ICD-10-PCS | 0PBG% | Excision / Humeral Shaft, Left |
| ICD-10-PCS | 0RBE% | Excision / Sternoclavicular Joint, Right |
| ICD-10-PCS | 0RBF% | Excision / Sternoclavicular Joint, Left |
| ICD-10-PCS | 0RBG% | Excision / Acromioclavicular Joint, Right |
| ICD-10-PCS | 0RBH% | Excision / Acromioclavicular Joint, Left |
| ICD-10-PCS | 0RBJ% | Excision / Shoulder Joint, Right |
| ICD-10-PCS | 0RBK% | Excision / Shoulder Joint, Left |
| ICD-10-PCS | 0PC5% | Extirpation / Scapula, Right |
| ICD-10-PCS | 0PC6% | Extirpation / Scapula, Left |
| ICD-10-PCS | 0PC7% | Extirpation / Glenoid Cavity, Right |
| ICD-10-PCS | 0PC8% | Extirpation / Glenoid Cavity, Left |
| ICD-10-PCS | 0PC9% | Extirpation / Clavicle, Right |
| ICD-10-PCS | 0PCB% | Extirpation / Clavicle, Left |
| ICD-10-PCS | 0PCC% | Extirpation / Humeral Head, Right |
| ICD-10-PCS | 0PCD% | Extirpation / Humeral Head, Left |
| ICD-10-PCS | 0PCF% | Extirpation / Humeral Shaft, Right |
| ICD-10-PCS | 0PCG% | Extirpation / Humeral Shaft, Left |
| ICD-10-PCS | 0RCE% | Extirpation / Sternoclavicular Joint, Right |
| ICD-10-PCS | 0RCF% | Extirpation / Sternoclavicular Joint, Left |
| ICD-10-PCS | 0RCG% | Extirpation / Acromioclavicular Joint, Right |
| ICD-10-PCS | 0RCH% | Extirpation / Acromioclavicular Joint, Left |
| ICD-10-PCS | 0RCJ% | Extirpation / Shoulder Joint, Right |
| ICD-10-PCS | 0RCK% | Extirpation / Shoulder Joint, Left |
| ICD-10-PCS | 0PD5% | Extraction / Scapula, Right |
| ICD-10-PCS | 0PD6% | Extraction / Scapula, Left |
| ICD-10-PCS | 0PD7% | Extraction / Glenoid Cavity, Right |
| ICD-10-PCS | 0PD8% | Extraction / Glenoid Cavity, Left |
| ICD-10-PCS | 0PD9% | Extraction / Clavicle, Right |
| ICD-10-PCS | 0PDB% | Extraction / Clavicle, Left |
| ICD-10-PCS | 0PDC% | Extraction / Humeral Head, Right |
| ICD-10-PCS | 0PDD% | Extraction / Humeral Head, Left |
| ICD-10-PCS | 0PDF% | Extraction / Humeral Shaft, Right |
| ICD-10-PCS | 0PDG% | Extraction / Humeral Shaft, Left |
| ICD-10-PCS | 0RGE% | Fusion / Sternoclavicular Joint, Right |
| ICD-10-PCS | 0RGF% | Fusion / Sternoclavicular Joint, Left |
| ICD-10-PCS | 0RGG% | Fusion / Acromioclavicular Joint, Right |
| ICD-10-PCS | 0RGH% | Fusion / Acromioclavicular Joint, Left |
| ICD-10-PCS | 0RGJ% | Fusion / Shoulder Joint, Right |
| ICD-10-PCS | 0RGK% | Fusion / Shoulder Joint, Left |
| ICD-10-PCS | 0RHC% | Insertion / Temporomandibular Joint, Right |
| ICD-10-PCS | 0RHD% | Insertion / Temporomandibular Joint, Left |
| ICD-10-PCS | 0RHE% | Insertion / Sternoclavicular Joint, Right |
| ICD-10-PCS | 0RHF% | Insertion / Sternoclavicular Joint, Left |
| ICD-10-PCS | 0RHG% | Insertion / Acromioclavicular Joint, Right |
| ICD-10-PCS | 0RHH% | Insertion / Acromioclavicular Joint, Left |
| ICD-10-PCS | 0RHJ% | Insertion / Shoulder Joint, Right |
| ICD-10-PCS | 0RHK% | Insertion / Shoulder Joint, Left |
| ICD-10-PCS | 0PH5% | Insertion / Scapula, Right |
| ICD-10-PCS | 0PH6% | Insertion / Scapula, Left |
| ICD-10-PCS | 0PH7% | Insertion / Glenoid Cavity, Right |
| ICD-10-PCS | 0PH8% | Insertion / Glenoid Cavity, Left |
| ICD-10-PCS | 0PH9% | Insertion / Clavicle, Right |
| ICD-10-PCS | 0PHB% | Insertion / Clavicle, Left |
| ICD-10-PCS | 0PHC% | Insertion / Humeral Head, Right |
| ICD-10-PCS | 0PHD% | Insertion / Humeral Head, Left |
| ICD-10-PCS | 0PHF% | Insertion / Humeral Shaft, Right |
| ICD-10-PCS | 0PHG% | Insertion / Humeral Shaft, Left |
| ICD-10-PCS | 0RJE% | Inspection / Sternoclavicular Joint, Right |
| ICD-10-PCS | 0RJF% | Inspection / Sternoclavicular Joint, Left |
| ICD-10-PCS | 0RJG% | Inspection / Acromioclavicular Joint, Right |
| ICD-10-PCS | 0RJH% | Inspection / Acromioclavicular Joint, Left |
| ICD-10-PCS | 0RJJ% | Inspection / Shoulder Joint, Right |
| ICD-10-PCS | 0RJK% | Inspection / Shoulder Joint, Left |
| ICD-10-PCS | 0PN5% | Release / Scapula, Right |
| ICD-10-PCS | 0PN6% | Release / Scapula, Left |
| ICD-10-PCS | 0PN7% | Release / Glenoid Cavity, Right |
| ICD-10-PCS | 0PN8% | Release / Glenoid Cavity, Left |
| ICD-10-PCS | 0PN9% | Release / Clavicle, Right |
| ICD-10-PCS | 0PNB% | Release / Clavicle, Left |
| ICD-10-PCS | 0PNC% | Release / Humeral Head, Right |
| ICD-10-PCS | 0PND% | Release / Humeral Head, Left |
| ICD-10-PCS | 0PNF% | Release / Humeral Shaft, Right |
| ICD-10-PCS | 0PNG% | Release / Humeral Shaft, Left |
| ICD-10-PCS | 0RNE% | Release / Sternoclavicular Joint, Right |
| ICD-10-PCS | 0RNF% | Release / Sternoclavicular Joint, Left |
| ICD-10-PCS | 0RNG% | Release / Acromioclavicular Joint, Right |
| ICD-10-PCS | 0RNH% | Release / Acromioclavicular Joint, Left |
| ICD-10-PCS | 0RNJ% | Release / Shoulder Joint, Right |
| ICD-10-PCS | 0RNK% | Release / Shoulder Joint, Left |
| ICD-10-PCS | 0RPE% | Removal / Sternoclavicular Joint, Right |
| ICD-10-PCS | 0RPF% | Removal / Sternoclavicular Joint, Left |
| ICD-10-PCS | 0RPG% | Removal / Acromioclavicular Joint, Right |
| ICD-10-PCS | 0RPH% | Removal / Acromioclavicular Joint, Left |
| ICD-10-PCS | 0RPJ% | Removal / Shoulder Joint, Right |
| ICD-10-PCS | 0RPK% | Removal / Shoulder Joint, Left |
| ICD-10-PCS | 0PP5% | Removal / Scapula, Right |
| ICD-10-PCS | 0PP6% | Removal / Scapula, Left |
| ICD-10-PCS | 0PP7% | Removal / Glenoid Cavity, Right |
| ICD-10-PCS | 0PP8% | Removal / Glenoid Cavity, Left |
| ICD-10-PCS | 0PP9% | Removal / Clavicle, Right |
| ICD-10-PCS | 0PPB% | Removal / Clavicle, Left |
| ICD-10-PCS | 0PPC% | Removal / Humeral Head, Right |
| ICD-10-PCS | 0PPD% | Removal / Humeral Head, Left |
| ICD-10-PCS | 0PPF% | Removal / Humeral Shaft, Right |
| ICD-10-PCS | 0PPG% | Removal / Humeral Shaft, Left |
| ICD-10-PCS | 0PQ5% | Repair / Scapula, Right |
| ICD-10-PCS | 0PQ6% | Repair / Scapula, Left |
| ICD-10-PCS | 0PQ7% | Repair / Glenoid Cavity, Right |
| ICD-10-PCS | 0PQ8% | Repair / Glenoid Cavity, Left |
| ICD-10-PCS | 0PQ9% | Repair / Clavicle, Right |
| ICD-10-PCS | 0PQB% | Repair / Clavicle, Left |
| ICD-10-PCS | 0PQC% | Repair / Humeral Head, Right |
| ICD-10-PCS | 0PQD% | Repair / Humeral Head, Left |
| ICD-10-PCS | 0PQF% | Repair / Humeral Shaft, Right |
| ICD-10-PCS | 0PQG% | Repair / Humeral Shaft, Left |
| ICD-10-PCS | 0RQE% | Repair / Sternoclavicular Joint, Right |
| ICD-10-PCS | 0RQF% | Repair / Sternoclavicular Joint, Left |
| ICD-10-PCS | 0RQG% | Repair / Acromioclavicular Joint, Right |
| ICD-10-PCS | 0RQH% | Repair / Acromioclavicular Joint, Left |
| ICD-10-PCS | 0RQJ% | Repair / Shoulder Joint, Right |
| ICD-10-PCS | 0RQK% | Repair / Shoulder Joint, Left |
| ICD-10-PCS | 0RRE% | Replacement / Sternoclavicular Joint, Right |
| ICD-10-PCS | 0RRF% | Replacement / Sternoclavicular Joint, Left |
| ICD-10-PCS | 0RRG% | Replacement / Acromioclavicular Joint, Right |
| ICD-10-PCS | 0RRH% | Replacement / Acromioclavicular Joint, Left |
| ICD-10-PCS | 0RRJ% | Replacement / Shoulder Joint, Right |
| ICD-10-PCS | 0RRK% | Replacement / Shoulder Joint, Left |
| ICD-10-PCS | 0PR5% | Replacement / Scapula, Right |
| ICD-10-PCS | 0PR6% | Replacement / Scapula, Left |
| ICD-10-PCS | 0PR7% | Replacement / Glenoid Cavity, Right |
| ICD-10-PCS | 0PR8% | Replacement / Glenoid Cavity, Left |
| ICD-10-PCS | 0PR9% | Replacement / Clavicle, Right |
| ICD-10-PCS | 0PRB% | Replacement / Clavicle, Left |
| ICD-10-PCS | 0PRC% | Replacement / Humeral Head, Right |
| ICD-10-PCS | 0PRD% | Replacement / Humeral Head, Left |
| ICD-10-PCS | 0PRF% | Replacement / Humeral Shaft, Right |
| ICD-10-PCS | 0PRG% | Replacement / Humeral Shaft, Left |
| ICD-10-PCS | 0RSE% | Reposition / Sternoclavicular Joint, Right |
| ICD-10-PCS | 0RSF% | Reposition / Sternoclavicular Joint, Left |
| ICD-10-PCS | 0RSG% | Reposition / Acromioclavicular Joint, Right |
| ICD-10-PCS | 0RSH% | Reposition / Acromioclavicular Joint, Left |
| ICD-10-PCS | 0RSJ% | Reposition / Shoulder Joint, Right |
| ICD-10-PCS | 0RSK% | Reposition / Shoulder Joint, Left |
| ICD-10-PCS | 0PS5% | Reposition / Scapula, Right |
| ICD-10-PCS | 0PS6% | Reposition / Scapula, Left |
| ICD-10-PCS | 0PS7% | Reposition / Glenoid Cavity, Right |
| ICD-10-PCS | 0PS8% | Reposition / Glenoid Cavity, Left |
| ICD-10-PCS | 0PS9% | Reposition / Clavicle, Right |
| ICD-10-PCS | 0PSB% | Reposition / Clavicle, Left |
| ICD-10-PCS | 0PSC% | Reposition / Humeral Head, Right |
| ICD-10-PCS | 0PSD% | Reposition / Humeral Head, Left |
| ICD-10-PCS | 0PSF% | Reposition / Humeral Shaft, Right |
| ICD-10-PCS | 0PSG% | Reposition / Humeral Shaft, Left |
| ICD-10-PCS | 0PT7% | Resection / Glenoid Cavity, Right |
| ICD-10-PCS | 0PT8% | Resection / Glenoid Cavity, Left |
| ICD-10-PCS | 0PT9% | Resection / Clavicle, Right |
| ICD-10-PCS | 0PTB% | Resection / Clavicle, Left |
| ICD-10-PCS | 0PTC% | Resection / Humeral Head, Right |
| ICD-10-PCS | 0PTD% | Resection / Humeral Head, Left |
| ICD-10-PCS | 0PTF% | Resection / Humeral Shaft, Right |
| ICD-10-PCS | 0PTG% | Resection / Humeral Shaft, Left |
| ICD-10-PCS | 0RTE% | Resection / Sternoclavicular Joint, Right |
| ICD-10-PCS | 0RTF% | Resection / Sternoclavicular Joint, Left |
| ICD-10-PCS | 0RTG% | Resection / Acromioclavicular Joint, Right |
| ICD-10-PCS | 0RTH% | Resection / Acromioclavicular Joint, Left |
| ICD-10-PCS | 0RTJ% | Resection / Shoulder Joint, Right |
| ICD-10-PCS | 0RTK% | Resection / Shoulder Joint, Left |
| ICD-10-PCS | 0RUE% | Supplement / Sternoclavicular Joint, Right |
| ICD-10-PCS | 0RUF% | Supplement / Sternoclavicular Joint, Left |
| ICD-10-PCS | 0RUG% | Supplement / Acromioclavicular Joint, Right |
| ICD-10-PCS | 0RUH% | Supplement / Acromioclavicular Joint, Left |
| ICD-10-PCS | 0RUJ% | Supplement / Shoulder Joint, Right |
| ICD-10-PCS | 0RUK% | Supplement / Shoulder Joint, Left |
| ICD-10-PCS | 0PU5% | Supplement / Scapula, Right |
| ICD-10-PCS | 0PU6% | Supplement / Scapula, Left |
| ICD-10-PCS | 0PU7% | Supplement / Glenoid Cavity, Right |
| ICD-10-PCS | 0PU8% | Supplement / Glenoid Cavity, Left |
| ICD-10-PCS | 0PU9% | Supplement / Clavicle, Right |
| ICD-10-PCS | 0PUB% | Supplement / Clavicle, Left |
| ICD-10-PCS | 0PUC% | Supplement / Humeral Head, Right |
| ICD-10-PCS | 0PUD% | Supplement / Humeral Head, Left |
| ICD-10-PCS | 0PUF% | Supplement / Humeral Shaft, Right |
| ICD-10-PCS | 0PUG% | Supplement / Humeral Shaft, Left |
| ICD-10-PCS | 0PW7% | Revision / Glenoid Cavity, Right |
| ICD-10-PCS | 0PW8% | Revision / Glenoid Cavity, Left |
| ICD-10-PCS | 0PW9% | Revision / Clavicle, Right |
| ICD-10-PCS | 0PWB% | Revision / Clavicle, Left |
| ICD-10-PCS | 0PWC% | Revision / Humeral Head, Right |
| ICD-10-PCS | 0PWD% | Revision / Humeral Head, Left |
| ICD-10-PCS | 0PWF% | Revision / Humeral Shaft, Right |
| ICD-10-PCS | 0PWG% | Revision / Humeral Shaft, Left |
| ICD-10-PCS | 0RWE% | Revision / Sternoclavicular Joint, Right |
| ICD-10-PCS | 0RWF% | Revision / Sternoclavicular Joint, Left |
| ICD-10-PCS | 0RWG% | Revision / Acromioclavicular Joint, Right |
| ICD-10-PCS | 0RWH% | Revision / Acromioclavicular Joint, Left |
| ICD-10-PCS | 0RWJ% | Revision / Shoulder Joint, Right |
| ICD-10-PCS | 0RWK% | Revision / Shoulder Joint, Left |
